# The potential health and economic impact of introducing the vaccine candidate VPM1002 to prevent tuberculosis disease in low- and middle-income countries: a modelling study

**DOI:** 10.64898/2026.08.17.26360583

**Authors:** Rebecca A Clark, Allison Portnoy, Tom Sumner, Daniel J Grint, Tomos O Prŷs-Jones, Roel Bakker, Nicolas A Menzies, Richard G White

## Abstract

**Background:** The tuberculosis (TB) vaccine candidate VPM1002 did not prove efficacy in the recent Phase III trial and is in discussion with the Indian regulator. However, low efficacy TB vaccines may still have public health value. We estimated the potential health and economic impact of introducing VPM1002 in low- and middle-income countries (LMICs).

**Methods:** We calibrated compartmental TB dynamic models to epidemiologic and demographic data for 79 individual LMICs. We assumed the vaccine would be introduced between 2027-2047, delivered routinely and annually to the age six cohort and delivered in two 10-yearly campaigns for older ages, be efficacious for 3 years, have efficacy of 16.9% (95% confidence interval = -13.3 to 39.1%), and prevent TB disease. We estimated the cumulative symptomatic TB episodes and TB-associated deaths averted by 2050, and cost-effectiveness from health-system and societal perspectives.

**Results:** Results suggest, across 79 LMICs, there may be 6.7 (95% uncertainty interval = -4.0 to 14.9) million symptomatic TB episodes averted, and 0.7 (-0.4 to 1.5) million TB-associated deaths averted overall over 2027-2050. At an assumed vaccine cost of 0.75 USD per dose, VPM1002 vaccination may be cost-effective compared to no vaccination in 15 of 79 modelled LMICs (19%), assuming a threshold of 1-times per-capita gross domestic product from the health system perspective, and may be cost-effective in 28 out of 79 countries (35%) and dominant in 14 countries (18%) from the societal perspective.

**Conclusions:** The VPM1002 Phase III trial did not prove efficacy, therefore results could be due to chance. However, if the true vaccine efficacy was consistent with the observed point estimate, then overall rollout in LMICs may avert a portion of symptomatic TB cases and TB-associated deaths, and in some countries could be cost-effective/saving. Although potentially infeasible, it would be useful to obtain more precise estimates of VPM1002 efficacy through larger Phase III/IV studies.

## INTRODUCTION

Tuberculosis (TB) is a serious global health problem, and a new TB vaccine for adolescents and adults could be an impactful strategy to reduce global burden.^1^ There is modelling evidence available on the potential impact for generic new TB vaccine product characteristics and the novel TB vaccine candidates M72/AS01_E_ and Vaccae,^2–6^ but limited evidence for any other specific candidates. Modelling suggests the most rapid and greatest impact would be from a new prevention of disease (POD) TB vaccine that is effective in uninfected and infected individuals and targeted to adolescents/adults.^7^ Modelling also suggests vaccines of this type with 50% efficacy and 10 years duration might avert 37–76 million cases and 5–8 million deaths in 105 low- and middle-income countries (LMICs) before 2050,^8,9^ and new TB vaccines with efficacy as low as 20% and protective duration as short as five years might also be cost-effective in some LMICs.^10^

The vaccine candidate VPM1002 is a recombinant urease C-deficient listeriolysin (Hly) expressing Bacillus Calmette-Guérin (BCG) vaccine strain sponsored by Serum Institute of India.^11^ A large Phase III three-arm POD trial of VPM1002 and Immuvac compared to placebo was recently completed in 12,717 household contacts aged 6 years and above of pulmonary TB (PTB) patients across India. The pre-specified primary endpoint of the trial was to evaluate vaccine efficacy by comparing the reduction in incidence of microbiologically-confirmed extrapulmonary (EPTB) and/or PTB over three years compared to placebo in the modified intention-to-treat (mITT) group.^12^ Results from the trial demonstrated no statistically significant impact against the primary endpoint, with an estimated efficacy of 16.9% (95% confidence interval = -13.3 to 39.1%),^13^ and is in discussion with the Indian regulator.^14^

The trial was powered to detect a vaccine efficacy of 50%. It may be possible that VPM1002 was efficacious at a lower efficacy but did not yield statistically significant results during this trial. It may also be that these results were due to chance.^15^ With these possibilities in mind, we used mathematical modelling to explore what the potential global impact of VPM1002 used as a POD vaccine might be, incorporating the uncertainty in efficacy observed in the trial.

## METHODS

### Study setting and data

We aimed to model countries classified as low- or middle-income as defined by the World Bank.^16^ Data to inform the country-specific TB incidence, mortality, and case notifications were from the World Health Organization (WHO) Global TB Report 2024.^1^ Data to inform the country-specific HIV prevalence and antiretroviral treatment (ART) coverage were from UNAIDS.^17^ The ratio of asymptomatic TB to all infectious TB was based on Frascella et al.^18^ Population estimates for each country from 1950–2100 were obtained from the United Nations Population Estimates and Projections, 2022 revision.^19^ Vaccine efficacy estimates and ranges were from Singh et al., 2026.^13^

### Model development and calibration

We used previously developed country-specific compartmental dynamic TB infection transmission models,^8,9,20^ updated to fit current TB burden estimates.^1^ We calibrated each country using history matching with emulation using the *hmer* R package until we had at least 200 parameter sets.^21^ We calibrated each country independently to eight country-specific TB targets: the TB incidence rate (all ages, ages 0–14, ages 15+);^1^ the TB mortality rate (all ages);^1^ the TB case notification rate (all ages, ages 0–14, ages 15+);^1^ and the global proportion of asymptomatic TB among infectious TB for all ages.^18^ For countries with a higher TB burden due to HIV, we additionally calibrated to at least three country-specific all-age targets: HIV prevalence, ART coverage, TB incidence rate in people living with HIV, and TB mortality rate in people living with HIV.^1,18^ We used the distribution of results produced by these parameter sets to quantify parameter uncertainty. Further details on model development and calibration are in Supplementary Material.

### No-new-vaccine baseline

Using the calibrated country-specific models, we simulated a no-new-vaccine baseline scenario for each country from 2010–2050 assuming the quality and coverage of non-vaccine interventions remained at 2023 levels, and no new TB vaccines are introduced. We did not explicitly model neonatal BCG vaccination the high level of reported coverage by countries is implicitly included in the calibration targets, and we do not anticipate it to be discontinued during the modelled time horizon.

### Vaccine scenarios

#### Main analysis

Based on trial results, we modelled a *Main analysis* scenario focused on the primary outcome of preventing microbiologically-confirmed PTB and/or EPTB using estimates for the mITT cohort (Table 1).^13^ We used the efficacy estimated for the primary endpoint for participants aged 6+ with any tuberculin skin test (TST) status at baseline (16.9% [-13.3 to 39.1]).^13^ We assumed no pre-vaccination infection status testing implying vaccine delivery to everyone within the target age group except for those with symptomatic TB or receiving TB treatment. We assumed the vaccine would be effective if delivered to those who were uninfected or infected at the time of vaccination. We assumed that everyone who was effectively vaccinated and protected would benefit from the vaccine by a reduction in the rate of progression to infectious disease (‘degree’ protection). We assumed the duration of protection would be three years on average with exponential waning.

**Table 1.** Modelled VPM1002 vaccine scenarios.

| Vaccine characteristics |  |  |  |  | Delivery characteristics |  |  |  |
| --- | --- | --- | --- | --- | --- | --- | --- | --- |
| Scenario<br>(author assigned names) | Efficacy<br>(95 % confidence<br>intervals)<sup>13</sup> | Durability | Who the<br>vaccine<br>works in | Vaccine<br>mechanism<br>of action | Age<br>targeting | Delivery | Coverage | Introduction<br>year |
| **Main analysis** | 16.9%<br>(-13.3 to 39.1) | 3 years | Uninfected<br>and infected | Prevents all<br>TB | 6+ | Annual routine to age 6, 10<br>yearly campaigns for age 7+ | Routine = 80%,<br>Campaign = 70% | Country-specific<br>years |
| **Varying duration of protection and introduction year scenarios** |  |  |  |  |  |  |  |  |
| **10 years duration of**<br>**protection** | 16.9%<br>(-13.3 to 39.1) | 10 years | Uninfected<br>and infected | Prevents all<br>TB | 6+ | Annual routine to age 6, 10<br>yearly campaigns for age 7+ | Routine = 80%,<br>Campaign = 70% | Country-specific<br>years |
| **All introduce in 2027** | 16.9%<br>(-13.3 to 39.1) | 3 years | Uninfected<br>and infected | Prevents all<br>TB | 6+ | Annual routine to age 6, 10<br>yearly campaigns for age 7+ | Routine = 80%,<br>Campaign = 70% | All introduce in<br>2027 |
| **Early adopters** | 16.9%<br>(-13.3 to 39.1) | 3 years | Uninfected<br>and infected | Prevents all<br>TB | 6+ | Annual routine to age 6, 10<br>yearly campaigns for age 7+ | Routine = 80%,<br>Campaign = 70% | Nine in 2026 or<br>country-specific |
| **Alternative efficacy scenarios** |  |  |  |  |  |  |  |  |
| **Per Protocol** | 21.4%<br>(-8.9 to 43.2) | 3 years | Uninfected<br>and infected | Prevents all<br>TB | 6+ | Annual routine to age 6, 10<br>yearly campaigns for age 7+ | Routine = 80%,<br>Campaign = 70% | Country-specific<br>years |
| **Prevents PTB-only** | 13.6%<br>(-21.1 to 38.3) | 3 years | Uninfected<br>and infected | Prevents<br>PTB-only | 6+ | Annual routine to age 6, 10<br>yearly campaigns for age 7+ | Routine = 80%,<br>Campaign = 70% | Country-specific<br>years |
| **Young** | 27.2%<br>(-14.8 to 53.9) | 3 years | Uninfected<br>and infected | Prevents all<br>TB | 6–18-year-olds | Annual routine to age 6, 10<br>yearly campaigns for age 7–18 | Routine = 80%,<br>Campaign = 70% | Country-specific<br>years |
| **Uninfected** | 39.0%<br>(-2.7 to 63.8) | 3 years | Uninfected<br>only | Prevents all<br>TB | 6+ | Annual routine to age 6, 10<br>yearly campaigns for age 7+ | Routine = 80%,<br>Campaign = 70% | Country-specific<br>years |

*Abbreviations: PTB = pulmonary tuberculosis, TB = tuberculosis.*

We assumed vaccine delivery routinely and annually to those aged six reaching 80% coverage over a five-year period, and two campaigns 10 years apart for ages seven and older reaching 70% coverage (over a five-year period for the first campaign and within one year for the second). We assumed delivery in country-specific introduction years as in the WHO investment case for new TB vaccines (shifted for earliest delivery in 2027, Table S6).^8^

#### Varying duration of protection and introduction years

We modelled three scenarios from the *Main Analysis* by varying the duration of protection from three years to ten years, the introduction year to all countries introducing in 2027, and the introduction year for nine countries anticipated to be early adopters to 2026 with country-specific years for the remaining 70. All other vaccine and delivery characteristics remained the same as the *Main analysis*.

#### Alternative efficacy scenarios

We modelled two different efficacy scenarios from the *Main Analysis*, using: (1) the per-protocol cohort estimate instead of the mITT cohort estimate (“*Per protocol”*) which estimated a vaccine efficacy of 21.4% (-8.9 to 43.2);^13^ and (2) the preventing PTB-only outcome instead of the preventing all TB outcome (“*Preventing PTB-only”*) which estimated a vaccine efficacy of 13.6% (-21.1 to 38.3).^13^ All other vaccine and delivery characteristics remained the same as the *Main analysis*.

The secondary endpoints suggested that there may be variation in efficacy by age group. We simulated the “*Young*” scenario, using the efficacy estimated for participants aged 6–18 year olds with any TST status at baseline (27.2% [-14.8 to 53.9]).^13^ We assumed the vaccine would be delivered routinely and annually to those aged six reaching 80% coverage over a five-year period, and as two 10-yearly campaigns for ages seven to eighteen reaching 70% coverage (over a five-year period for the first campaign and over one year for the second). Compared to the *Main analysis*, we changed both the efficacy and ages targeted, while all other vaccine and delivery characteristics remained the same.

Finally, we assumed the vaccine would only be effective if delivered to those who were uninfected at the time of vaccination, and that those who were infected at the time of vaccination would not receive any benefit from the vaccine— the “*Uninfected*” scenario, using the efficacy estimated for the sub-group of participants aged six and over who were TST-at baseline (39.0% [-2.7 to 63.8]).^13^ Compared to the *Main analysis*, we changed both the efficacy and who the vaccine was assumed to work in, while all other vaccine and delivery characteristics remained the same.

Due to considerable uncertainty in efficacy estimates, often with negative efficacy as a lower bound, instead of using point estimates, common in modelling studies, we simulated the distribution for each empirical vaccine efficacy range using the confidence intervals reported from the trial. For each vaccine scenario simulation, we matched a unique parameter set with a unique efficacy estimate from the distribution to estimate population-level impact. We reported the median and the 95% uncertainty range from 200 simulations.

### Health outcomes

For each vaccine scenario, we calculated the incidence and mortality rate reductions in 2050, and the cumulative number of symptomatic TB episodes and TB-associated deaths averted between vaccine introduction and 2050 compared to the no-new-vaccine baseline. We estimated disability-adjusted life-years (DALYs) averted, calculated as the summed years of healthy life lost due to disability (YLDs) and years of healthy life lost due to premature death (YLLs), to quantify the health gains achieved by vaccination in the cost-effectiveness analysis. To calculate YLDs, we assigned each modelled health state a disability weight from the Global Burden of Disease classification system and summed life years lived across all health states, weighted by the disability weight for each state.^21^ To calculate YLLs, we multiplied deaths at each year of age by reference life expectancy at that age and summing across all ages.^22^

### Cost outcomes

We estimated the costs of vaccine introduction and changes in costs of other health services (TB care, HIV care) by multiplying health service volume indicators (vaccines delivered, TB cases diagnosed and treated, ART patient-years) by country-specific unit costs. Country-specific unit costs for diagnostic costs, TB treatment costs, ART costs, vaccine delivery costs, vaccine introduction costs, direct non-medical costs (travel, accommodation, food, nutritional supplements) to the patient, and income losses experienced by patients during TB care have been previously described.^19^ Productivity costs due to premature death were estimated as the incremental number of life-years gained under a given vaccination scenario, multiplied by 2023 per-capita gross domestic product (GDP) as an approximation of income.

The base-case vaccine price for VPM1002 was assumed to be $0.75 per dose with an injection supply cost per dose of $0.11 and 5% wastage. Costs are reported in 2023 US dollars.

### Cost-effectiveness analysis

Incremental cost-effectiveness ratios (ICERs) were calculated from health system and societal perspectives, with a 3% discount rate, across the 2026 to 2050 evaluation period. The health system perspective considered costs of vaccine introduction, plus the costs of TB and HIV services indirectly affected by vaccine introduction. The societal perspective additionally included patient non-medical and productivity costs. ICERs were compared to a range of country-specific cost-effectiveness thresholds as multiples of per-capita GDP (assuming 1× per-capita GDP as a proxy for willingness to pay in the base-case) to reflect the lack of consensus for a single threshold.^25^ We identified introduction of VPM1002 as ‘dominant’ if it was found to be both less costly and more effective (in this analysis, greater DALYs averted) compared to no-new-vaccine introduction, whereas we identified introduction of VPM1002 to be ‘dominated’ if it was found to be both more costly and less effective compared to no-new-vaccine introduction.

### Additional price scenarios

We modelled two alternative vaccine price scenarios assuming $0.50 and $1.00 per dose, respectively.

## RESULTS

We included results for 79 LMICs, accounting for 91% of global TB incidence and 90% of global TB mortality in 2024. Incorporating the observed uncertainty in efficacy from the trial, across the 79 countries modelled, the *Main analysis* scenario may avert 6.7 (95% uncertainty interval = -4.0 to 14.9) million symptomatic TB episodes and 0.7 (-0.4 to 1.5) million TB-associated deaths over 2027–2050 (Table 2, Figure 1). The uncertainty range included some simulations (10% in the *Main analysis*) in which there was more TB with vaccination than without, in line with the efficacy confidence interval from the trial, which crossed zero.

**Table 2.** Epidemiological and economic outcomes of the VPM1002 candidate TB vaccine by analytic scenario.

| Scenario | Efficacy<sup>13</sup> | Percent incidence rate reduction in 2050 (%) | Percent mortality rate reduction in 2050 (%) | Cumulative sTB episodes averted by 2050 (millions) | Cumulative TB deaths averted by 2050 (millions) | Health system perspective<sup>a</sup> incremental cost (USD billions) | Societal perspective<sup>b</sup> incremental cost (USD billions) | DALYs averted (millions) | Health system cost (USD) per DALY averted | Societal cost (USD) per DALY averted |
| --- | --- | --- | --- | --- | --- | --- | --- | --- | --- | --- |
| **Main analysis** | 16.9%<br>(-13.3 to 39.1) | 4.1%<br>(-2.4 to 9.2) | 4.8%<br>(-2.8 to 10.9) | 6.7<br>(-4.0 to 14.9) | 0.7<br>(-0.4 to 1.5) | 40.5<br>(32.3 to 56.0) | 11.4<br>(-31.6 to 54.8) | 11.0<br>(-6.9 to 26.4) | 3676<br>(1343 to dominated) | 1037<br>(dominant to dominated) |
| **Varying duration of protection and introduction year scenarios** |  |  |  |  |  |  |  |  |  |  |
| **10 years duration of protection** | 16.9%<br>(-13.3 to 39.1) | 9.3%<br>(-5.5 to 20.1) | 10.0%<br>(-6.1 to 22.2) | 12.2<br>(-7.5 to 26.7) | 1.2<br>(-0.7 to 2.6) | 34.6<br>(27.2 to 47.9) | -15.3<br>(-85.7 to 63.9) | 18.9<br>(-12.3 to 44.5) | 1830<br>(660 to dominated) | dominant<br>(dominant to dominated) |
| **All introduce in 2027** | 16.9%<br>(-13.3 to 39.1) | 3.5%<br>(-2.0 to 8.0) | 4.1%<br>(-2.4 to 9.4) | 8.1<br>(-4.8 to 18.3) | 0.9<br>(-0.5 to 2.0) | 43.3<br>(34.3 to 59) | 3.2<br>(-54.4 to 64.4) | 15.1<br>(-9.3 to 35.8) | 2875<br>(1066 to dominated) | 215<br>(dominant to dominated) |
| **Early adopters** | 16.9%<br>(-13.3 to 39.1) | 3.6%<br>(-2.1 to 8.3) | 4.2%<br>(-2.4 to 9.7) | 7.8<br>(-4.6 to 17.5) | 0.9<br>(-0.5 to 1.9) | 42.2<br>(33.3 to 57.8) | 2.8<br>(-54.7 to 63) | 14.4<br>(-8.7 to 34.4) | 2939<br>(1108 to dominated) | 196<br>(dominant to dominated) |
| **Alternative efficacy scenarios** |  |  |  |  |  |  |  |  |  |  |
| **Per protocol** | 21.4%<br>(-8.9 to 43.2) | 5.0%<br>(-0.6 to 10.4) | 6.0%<br>(-0.8 to 11.9) | 8.2<br>(-1.0 to 16.5) | 0.8<br>(-0.1 to 1.7) | 40.3<br>(32.7 to 54.9) | 2.9<br>(-42.5 to 50.6) | 14.1<br>(-1.9 to 30.3) | 2849<br>(1258 to dominated) | 203<br>(dominant to dominated) |
| **Prevents PTB-only** | 13.6%<br>(-21.1 to 38.3) | 3.2%<br>(-4.6 to 8.3) | 3.8%<br>(-5.5 to 10.0) | 5.1<br>(-6.5 to 14.2) | 0.5<br>(-0.7 to 1.4) | 40.7<br>(32.4 to 55.8) | 19.1<br>(-25.6 to 73.5) | 8.2<br>(-11.6 to 23.2) | 4986<br>(1554 to dominated) | 2340<br>(dominant to dominated) |
| **Young** | 27.2%<br>(-14.8 to 53.9) | 3.1%<br>(-2.3 to 6.8) | 3.7%<br>(-2.7 to 8.1) | 4.3<br>(-3.1 to 9.2) | 0.5<br>(-0.4 to 1.0) | 9.9<br>(8.4 to 12.1) | -11.4<br>(-39.8 to 25.7) | 8.3<br>(-6.57 to 19.2) | 1200<br>(477 to dominated) | dominant<br>(dominant to dominated) |
| **Uninfected** | 39.0%<br>(-2.7 to 63.8) | 4.7%<br>(-0.6 to 8.1) | 5.4%<br>(-0.7 to 9.7) | 6.5<br>(-0.8 to 12.1) | 0.7<br>(-0.1 to 1.2) | 42.5<br>(34.4 to 59) | 14.9<br>(-17.1 to 56.9) | 10.6<br>(-1.4 to 20.5) | 4010<br>(1798 to dominated) | 1406<br>(dominant to dominated) |

<sup>a</sup> Costs from the health system perspective include vaccination costs, TB testing and treatment costs, and antiretroviral treatment costs,
<sup>b</sup> Costs from the societal perspective include health system perspective costs, as well as patient non-medical costs and productivity losses.
Note: Introduction of VPM1002 is considered 'dominant' if it is both less costly and more effective (in this analysis, greater DALYs averted) compared to no-new-vaccine introduction, whereas introduction of VPM1002 is considered 'dominated' if it is both more costly and less effective compared to no-new-vaccine introduction.
Abbreviations: DALY = disability-adjusted life year, PTB = pulmonary tuberculosis, sTB = symptomatic tuberculosis, TB = tuberculosis.

**Figure 1.**
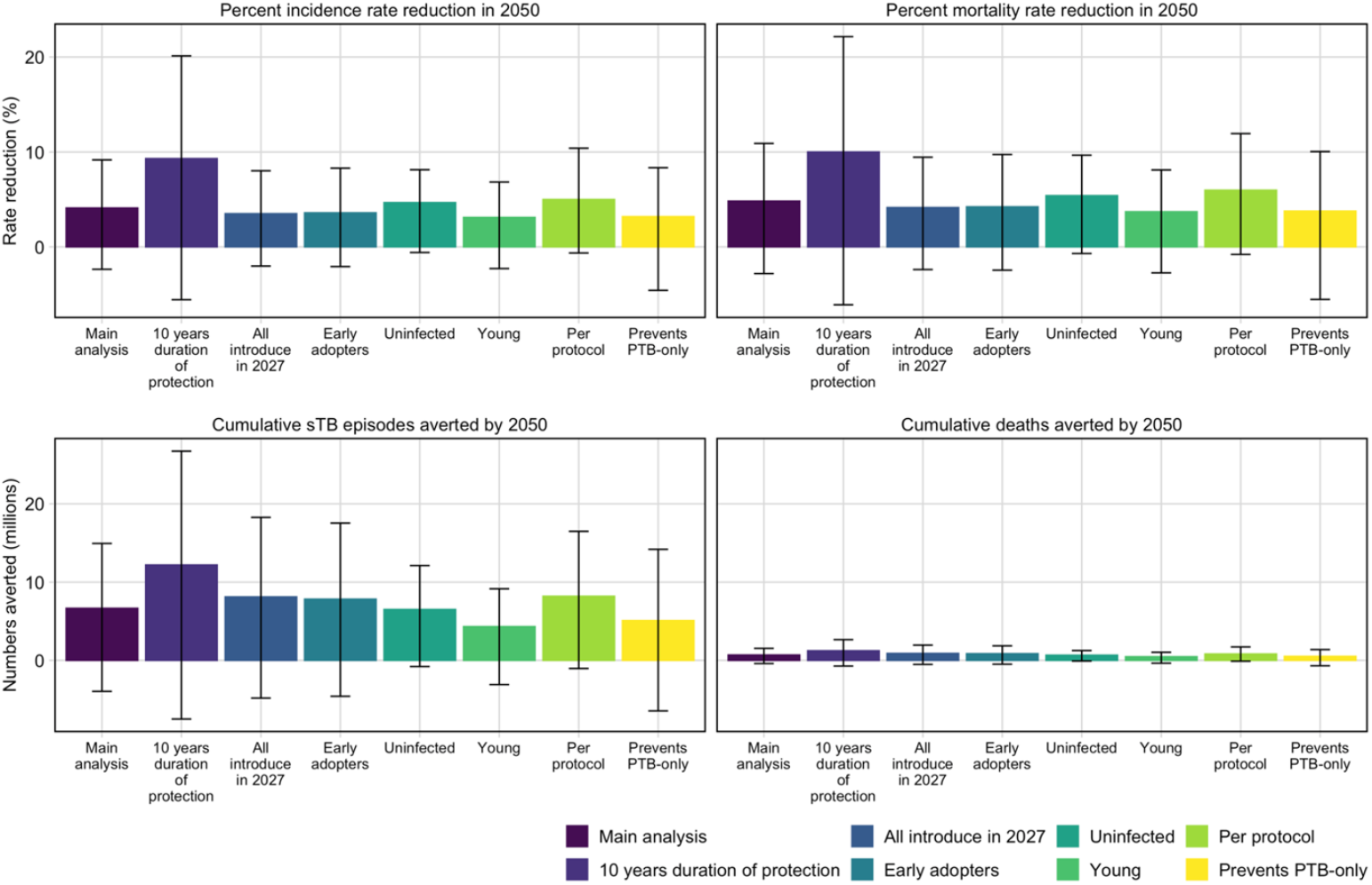
Incidence and mortality rate reductions in 2050, and cumulative number of symptomatic TB episodes and TB-associated deaths averted between vaccine introduction and 2050 for the main analysis compared to the alternative scenarios

Increasing the assumed average duration of protection from three years to ten years increased the magnitude of the potential symptomatic TB episodes averted to 12.2 (-7.5 to 26.7) million, and the potential TB-associated deaths averted to 1.2 (-0.7 to 2.6) million (Table 2, Figure 1). If all countries (unrealistically) introduced the vaccine in 2027 instead of over country-specific years, there was an increase in the magnitude of the potential symptomatic TB episodes and TB-associated deaths averted, but a limited change in incidence and mortality rate reductions in 2050 due to waning protection (Table 2, Figure 1). If instead the vaccine was introduced in 2026 for nine potential early adopter countries, we estimated 7.8 (-4.6 to 17.5) million symptomatic TB episodes and 0.9 (-0.5 to 1.9) million TB-associated deaths could be averted between 2026–2050 (Table 2, Figure 1).

The efficacy range was slightly higher in the *Per protocol* scenario, leading to the *Per protocol* scenario potentially averting 8.1 (-1.0 to 16.5) million symptomatic TB episodes and 0.8 (-0.1 to 1.7) million TB-associated deaths between 2027–2050 (Table 2, Figure 1). For the *Prevents PTB-only* scenario, the efficacy range was lower, and we estimated 5.1 (-6.5 to 14.2) million symptomatic TB episodes, and 0.5 (-0.7 to 1.4) million TB-associated deaths may be averted between 2027–2050 (Table 2, Figure 1). The *Young* and *Uninfected* scenarios could avert 4.3 (-3.1 to 9.2) million and 6.5 (-0.8 to 12.1) million symptomatic TB episodes, and 0.5 (-0.4 to 1.0) million and 0.7 (-0.1 to 1.2) million TB-associated deaths, respectively, between 2027–2050 (Table 2, Figure 1).

Table 2 reports summary health outcomes, costs, and cost-effectiveness of VPM1002 vaccination across all 79 modelled LMICs by scenario, for both the health system and societal perspectives. In the *Main analysis*, across 200 simulations, the health system perspective indicated uniformly higher costs, while the societal perspective often resulted in lower and frequently dominant outcomes, with many simulations falling in the southeast quadrant of the cost-effectiveness plane (Figure S 6). The probability that

VPM1002 was cost-effective increased with higher willingness-to-pay thresholds under both perspectives, with consistently greater probabilities observed from the societal perspective compared with the health system perspective (Figure S 7). In the *Main analysis*, from the health system perspective, we found that VPM1002 vaccination may be cost-effective (ICER below 1-times per-capita GDP) compared to no vaccination in 15 of 79 modelled LMICs (19%). From the societal perspective, VPM1002 may be cost-effective in 28 out of 79 countries (35%), and dominant in 14 countries (18%). For all scenarios, due to a negative health gain (i.e., negative DALYs averted) for the lower bound, introducing VPM1002 vaccine may be dominated compared to no introduction of a new vaccine for the upper bound of the uncertainty interval.

Figures 2 and 3 display the percentage of countries where vaccination may be cost-effective based on different cost-effectiveness thresholds, for the health system and societal perspectives, comparing the *Main analysis* to alternative scenarios. When considering increased duration of protection, accelerated vaccine introduction, or early adoption in nine countries, we found that the *Main analysis* may be cost-effective in more settings and with lower ICERs. VPM1002 vaccination may be cost-effective for 28 of 79 countries (35%) from the health system perspective and 42 of 79 countries (53%) from the societal perspective (including dominant in 29 countries) when assuming an increased duration of protection, for 21 of 79 countries (27%) from the health system perspective and 35 of 79 countries (44%) from the societal perspective (including dominant in 20 countries) when assuming an accelerated vaccine introduction timeline, and for 18 of 79 countries (23%) from the health system perspective and 29 of 79 countries (37%) from the societal perspective (including dominant in 16 countries) when assuming the specified nine countries introduce in 2026 (Figure 2). Alternative vaccine efficacy assumptions may improve both the magnitude of cost-effectiveness and the dominants by scenario (Figure 3).

**Figure 2.**
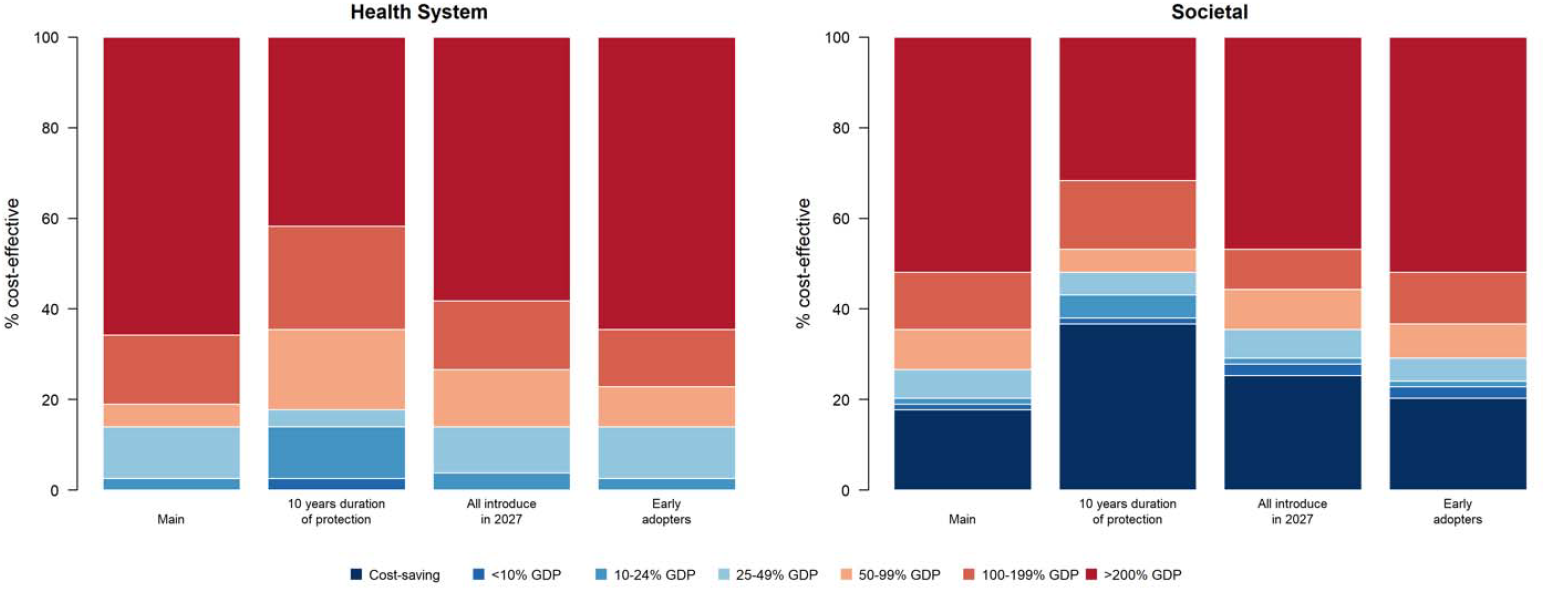
Percentage of countries where vaccination may be cost-effective compared to percentage of gross domestic product per capita thresholds, comparing health system and societal perspectives, for the *Main analysis* compared to 10 years duration of protection, all countries introducing in 2027, and early adopter countries, assuming $0.75 vaccine price per dose. Note: Countries include 79 low- and middle-income countries analysed. GDP per capita estimates from 2023. GDP, gross domestic product per capita.

**Figure 3.**
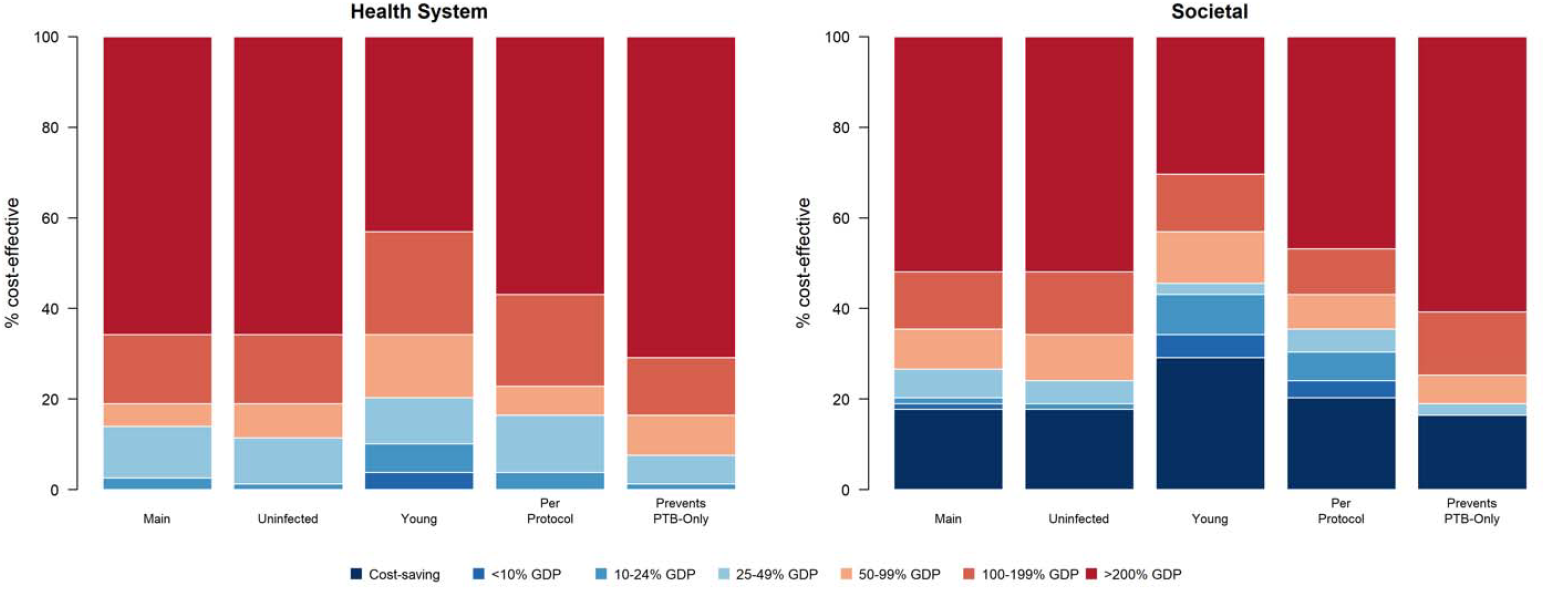
Percentage of countries where vaccination may be cost-effective compared to percentage of gross domestic product per capita thresholds, comparing health system and societal perspectives, by vaccine efficacy scenario, assuming $0.75 vaccine price per dose. Note: Countries include 79 low- and middle-income countries analysed. GDP per capita estimates from 2023. GDP, gross domestic product per capita.

Whether decreasing the vaccine price per dose to $0.50 (Table S8, Figures S8–S9) or increasing the vaccine price per dose to $1.00 (Table S9, Figures S10–S11, we estimated similar trends in terms of cost-effectiveness and dominants by scenario compared to the base-case assumption of $0.75 per dose.

However, as expected, estimated ICERs were lower in magnitude (more favourable) when assuming a vaccine price of $0.50 per dose and higher in magnitude when assuming a vaccine price of $1.00 per dose.

## DISCUSSION

Results suggest that, across 79 LMICs, introduction of the VPM1002 vaccine (with trial-reported efficacy) may avert 6.7 (-4.0 to 14.9) million symptomatic TB episodes averted and 0.7 (-0.4 to 1.5) million TB-associated deaths averted between 2027-2050. At an assumed vaccine cost of 0.75 USD per dose, VPM1002 vaccination may be cost-effective compared to no vaccination in 15 of 79 modelled LMICs (19%), assuming a threshold of 1-times per-capita gross domestic product from the health system perspective, and may be cost-effective in 28 out of 79 countries (35%) and dominant (both less costly and more effective compared to no-new-vaccine introduction) in 14 countries (18%) from the societal perspective.

The primary trial endpoint was not proven, and therefore these findings may be due to chance.^13^ Results from modelling a vaccine with efficacy aligned with the point estimate may be misleading about the impact possible from introducing VPM1002, given the uncertainty crosses zero, could increase the risk of TB or could have no impact at all, and implementation would use resources that may have greater impact if allocated to other TB prevention strategies. We ran scenarios using the range of lower and upper confidence limits from the trial reported hazard ratio to appropriately account for the uncertainty in the vaccine efficacy estimates and therefore results from some simulations led to an increase in the number of symptomatic TB episodes and TB-associated deaths by 2050. Similarly, while we found that introduction of VPM1002 could be considered cost-effective on average, it was dominated (both more costly and less effective) compared to no introduction on the lower bound of the uncertainty for all populations and scenarios of vaccine efficacy.

Compared to modelling of a hypothetical vaccine with 50% efficacy and 10 years protection, the introduction of VPM1002 would have a comparatively small impact.^8^ However, depending on the price of the vaccine, and the true vaccine efficacy, VPM1002 might still be a cost-effective intervention, even at low efficacy and if a more efficacious vaccine were not available. The estimated cost per dose of VPM1002 was USD $0.75 ($0.50 to $1.00), which is substantially lower than the previous estimates used for the vaccine during the WHO investment case for new TB vaccines (∼USD $4.60 per dose).

Previously, vaccines with lower efficacies, if proven in trials and acceptable to countries, have received recommendations for introduction if other conditions are met and may have a meaningful impact. For example, the RTS,S malaria vaccine was recommended by WHO in 2021 as it was deemed safe, feasible to deliver, and highly cost effective, while the trial report efficacy against severe malaria for four doses delivered to infants and children aged 5–17 months was 32.2% (95% confidence interval = 13.7 to 46.9).^26,27^

This was a mathematical modelling study, and therefore limitations associated with mathematical modelling apply. Model misspecification could be possible, as we used one consistent TB and HIV/ART model structure for all countries, calibrated to country-specific TB targets and represented uncertainty in the underlying natural history of TB. The list of LMICs from the World Bank includes 132 countries, however data was missing to attempt calibration for several countries, and several countries did not successfully calibrate. Excluding these countries will reduce the number of averted symptomatic TB episodes and TB-associated deaths and could bias the generalisability of the relative impacts if the countries excluded were systemically different than those included. We assumed that non-vaccine interventions would continue at their 2023 coverage and effectiveness. This may not be the case; reductions to overseas funding assistance to support global health or other global shocks could cause the slowly declining trends to reverse, or new diagnostics or treatments could accelerate the declining trend. In these situations, we would have either under- or over-estimated the amount of TB in the population for a vaccine to avert. We focussed on scenarios where the vaccine prevented PTB disease, whereas post-hoc analysis of the trial data indicated there may be an effect for EPTB which was not evaluated in this study, but has been evaluated elsewhere.^28^

For the costing, we extrapolated from published literature for major unit cost inputs, as previously described in Portnoy et al. (2023).^20^ Therefore, the resulting cost estimates may not capture the full level of variation in these costs. In this analysis, we used 1-times per-capita GDP as our base-case cost-effectiveness threshold to approximate willingness-to-pay per DALY averted. However, there have been recent proposals that a more stringent cost-effectiveness threshold may be appropriate in LMIC settings,^29,30^ which could result in VPM1002 introduction being cost-effective in fewer countries.

The global burden of TB is substantial, and a new TB vaccine is likely needed to help reach elimination. If significant efficacy is confirmed for VPM1002, it may reduce symptomatic TB episodes and TB-associated deaths in LMICs and may be cost-effective, especially in high burden countries. It would be valuable to carry out larger or longer studies to provide more precise estimates of the efficacy of VPM1002, given the wide confidence intervals reported by the trial. Repeat or larger Phase III studies are needed and would need to be powered appropriately to detect lower efficacy than 50%.

## Data Availability

The analytic code used in this study will be made publicly available from the time of publication from https://github.com/lshtm-tbmg/VPM1002.

## Ethics approval

No ethical approval was needed for this study. All data used was obtained from publicly available sources.

