## Supplementary Material for "The potential health and economic impact of introducing the vaccine candidate VPM1002 to prevent tuberculosis disease in low- and middle-income countries: a modelling study"

**Supplementary Methods**

### 1. Model structure and parameterisation

We created a compartmental tuberculosis (TB) model which includes separate structures to account for key modelling components required, including age, TB natural history, HIV and ART, identical to that included in Clark et al.^1^ The information in the Clark et al., supplementary material is replicated here with minor modifications.

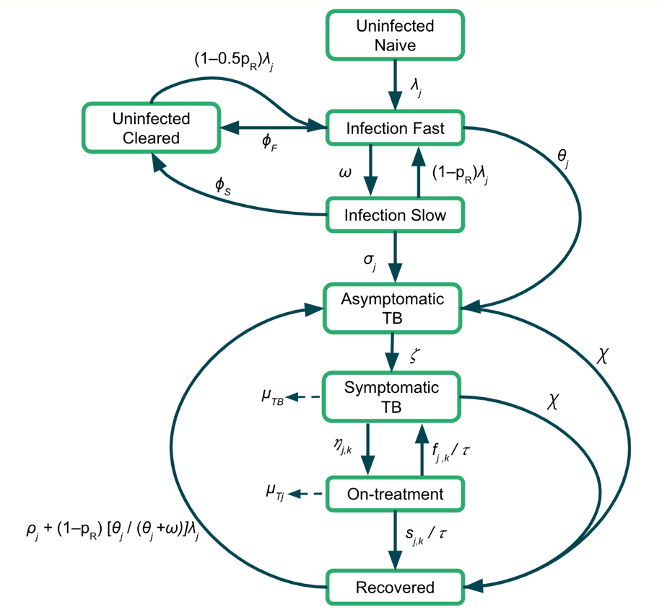

Figure S 1	TB natural history model structure*.*

*Subscript j represents parameters that vary by age, and subscript k represents parameters that vary over time.*

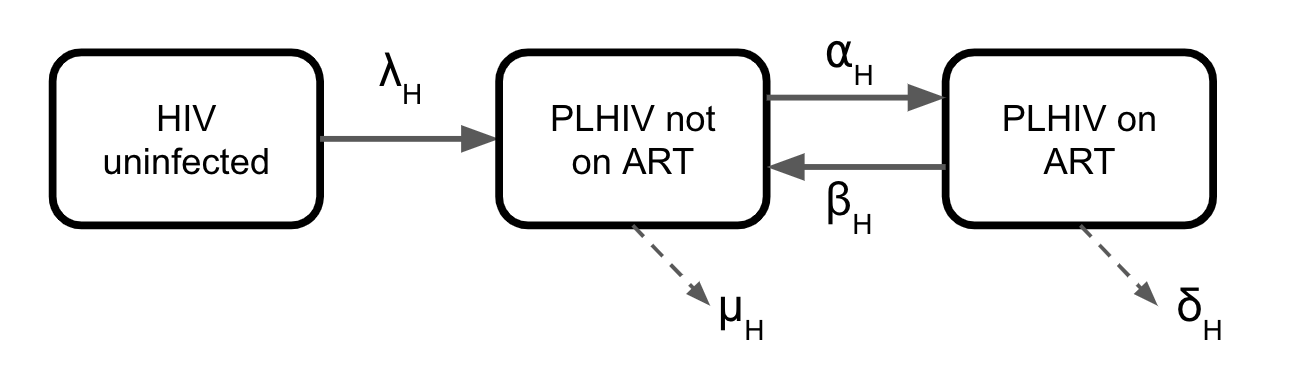

Figure S 2	HIV and ART structure

Parameters used in the natural history model structure and the HIV and ART model structure are provided in Table S 1 below, along with their definitions, sources, and information on whether the parameter is fixed or varied (as well as whether they are varied by age or time) during calibration. The parameter ranges provided for the tuberculosis natural history parameters are priors fitted during calibration in a Bayesian analysis. We assumed that all values within the prior range are equally likely. The prior ranges were pre-specified based on literature review and were reviewed as new data became available.

Table S 1	Demographic and TB natural history parameters and definitions.

| **Description** | **Units** | **Symbol** | **Prior** | **Fixed or varying** | **Age varying** | **Time varying** | **Source** |
| --- | --- | --- | --- | --- | --- | --- | --- |
| Birth rate | Per year | 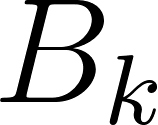<https://www.codecogs.com/eqnedit.php?latex=B_k#0> | United Nations population estimates and projections | Fixed | No | Yes | ^2^ |
| Background mortality rate | Per year | 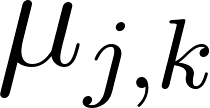<https://www.codecogs.com/eqnedit.php?latex=%5Cmu_%7Bj%2Ck%7D#0> | Calculated from United Nations population estimates and projections | Fixed | Yes | Yes | ^2^ |
| Mortality rate for sTB | Per person  per year | 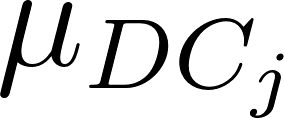<https://www.codecogs.com/eqnedit.php?latex=%5Cmu_%7BDC_j%7D#0> | (0–0.178) | Varying | Yes | No | ^3^ |
| Force of infection | Per year | 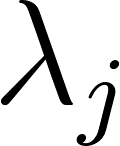<https://www.codecogs.com/eqnedit.php?latex=%5Clambda_j#0> | Fitted | Fixed equation | Yes | No | *Calculated* |
| Probability of transmission per infectious contact | - | 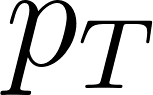<https://www.codecogs.com/eqnedit.php?latex=p_T#0> | (0–0.0068) | Varying | No | No | *Assumed* |
| Fraction of total TB disease that is EPTB | - | 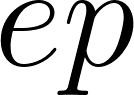<https://www.codecogs.com/eqnedit.php?latex=ep#0> | Country-specific average of previous 3 years | Fixed | No | No | ^4^ |
| Infectiousness of aTB relative to sTB | - | 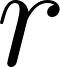<https://www.codecogs.com/eqnedit.php?latex=r#0> | 0.80 | Fixed | No | No | ^5^ |
| Rate of self-clearance from If to Uc | Per person per year | 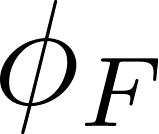<https://www.codecogs.com/eqnedit.php?latex=%5Cphi_F#0> | 0.00000140 | Fixed | No | No | ^6^ |
| Rate of self-clearance from Is to Uc | Per person per year | 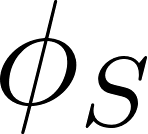<https://www.codecogs.com/eqnedit.php?latex=%5Cphi_S#0> | (0.0254–0.0467) | Varying | No | No | ^6^ |
| Rate of fast progression to aTB | Per person per year | 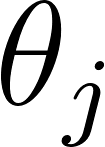<https://www.codecogs.com/eqnedit.php?latex=%5Ctheta_j#0> | (0.0696–0.111) | Varying | Yes | No | ^6^ |
| Rate from If to Is | Per person  per year | 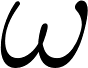<https://www.codecogs.com/eqnedit.php?latex=%5Comega#0> | 0.5 | Fixed | No | No | *Assumed* |
| Rate of reactivation  from Is | Per person  per year | 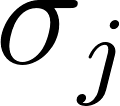<https://www.codecogs.com/eqnedit.php?latex=%5Csigma_j#0> | (0.000135–0.00113) | Varying | Yes | No | ^6^ |
| Rate of progression from aTB to sTB | Per person  per year | 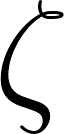<https://www.codecogs.com/eqnedit.php?latex=%20%5Czeta%20#0> | (0–12) | Varying | No | No | *Assumed* |
| Rate of natural cure from aTB and sTB | Per person  per year | 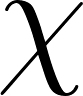<https://www.codecogs.com/eqnedit.php?latex=%5Cchi#0> | (0.1–0.25) | Varying | No | No | ^7,8^ |
| Rate of relapse from R | Per person per year | 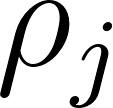<https://www.codecogs.com/eqnedit.php?latex=%5Crho_j#0> | (0.0001–0.07) | Varying | Yes | No | ^9,10^ |
| Protection from reinfection for If, Is, and R | - | 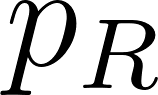<https://www.codecogs.com/eqnedit.php?latex=%20p_R%20#0> | (0.6–0.85) | Varying | No | No | ^8,11,12^ |
| ***HIV parameters*** | | | | | | | |
| HIV incidence rate fitting factor | - | 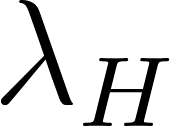<https://www.codecogs.com/eqnedit.php?latex=%5Clambda_H#0> | (0–300) | Varying | No | No | *Fitted* |
| Rate of ART initiation fitting factor | - | 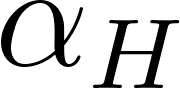<https://www.codecogs.com/eqnedit.php?latex=%5Calpha_H#0> | (0–7000) | Varying | No | No | *Fitted* |
| Rate of ART discontinuation | Per year | 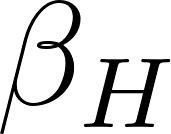<https://www.codecogs.com/eqnedit.php?latex=%5Cbeta_H#0> | 0.074 | Fixed | No | No | ^13,14^ |
| Mortality rate from  PLHIV not on ART | Per year | 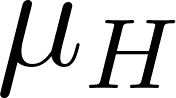<https://www.codecogs.com/eqnedit.php?latex=%5Cmu_H#0> | 0.10 | Fixed | No | No | ^15^ |
| Mortality rate from  PLHIV on ART | Per year | 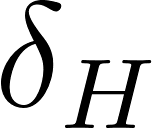<https://www.codecogs.com/eqnedit.php?latex=%5Cdelta_H#0> | 0.026 | Fixed | No | No | ^16^ |
| Relative increase in progression rate for PLHIV not on ART | - | 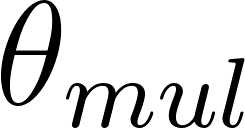<https://www.codecogs.com/eqnedit.php?latex=%5Ctheta_%7Bmul%7D#0> | (3.94–14.45) | Varying | No | No | ^17^ |
| Relative reduction in  for HIV and ART compartments | - | 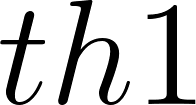<https://www.codecogs.com/eqnedit.php?latex=th1#0> | HIV uninfected = 0  PLHIV not on ART = 1.00  PLHIV on ART= 0.35 | Fixed | No | No | ^18^ |
| Relative mortality rate adjustment for HIV and ART compartments | - | 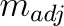 | HIV uninfected = 1.00  PLHIV not on ART = 1.50  PLHIV on ART = 1.15 | Fixed | No | No | ^3, 18–20^ |

Table S 2	Age varying parameters in the TB natural history model

| **Parameter** | **Range of Parameter** | **Age Varying Description** | **Age Scaling Parameter** | **Adults**  **(**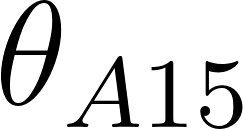<https://www.codecogs.com/eqnedit.php?latex=%5Ctheta_%7BA15%7D#0>**)** | **Children**  **(**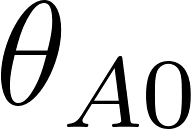<https://www.codecogs.com/eqnedit.php?latex=%5Ctheta_%7BA0%7D#0>**)** |
| --- | --- | --- | --- | --- | --- |
| 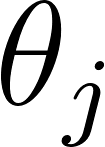<https://www.codecogs.com/eqnedit.php?latex=%5Ctheta_j#0>  Rate per year of fast progression | 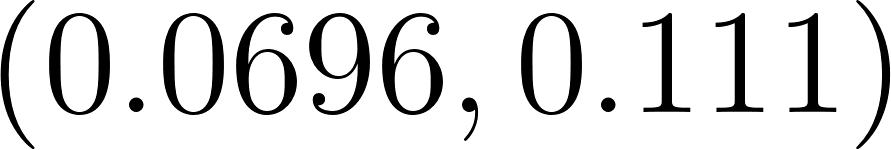<https://www.codecogs.com/eqnedit.php?latex=(0.0696%2C%200.111)#0> | Retain if value for children is **less** than value for adults | Sample 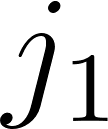<https://www.codecogs.com/eqnedit.php?latex=j_1#0> from 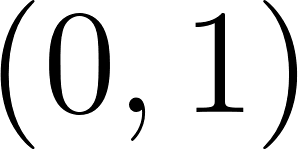<https://www.codecogs.com/eqnedit.php?latex=(0%2C1)#0> | Sample 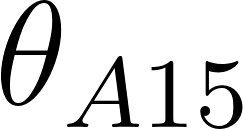<https://www.codecogs.com/eqnedit.php?latex=%5Ctheta_%7BA15%7D#0> from  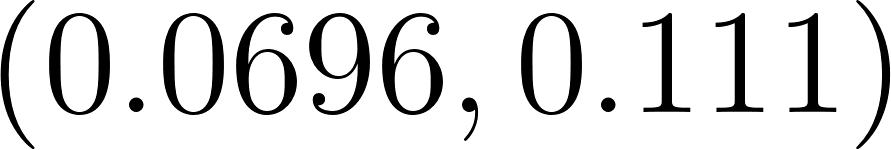<https://www.codecogs.com/eqnedit.php?latex=%20(0.0696%2C%200.111)#0> | 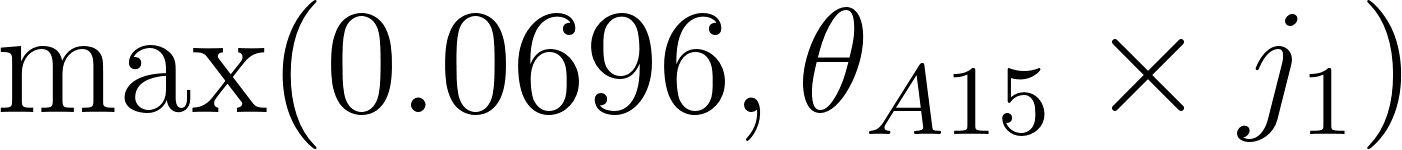<https://www.codecogs.com/eqnedit.php?latex=%5Cmax(0.0696%2C%20%5Ctheta_%7BA15%7D%20%5Ctimes%20j_1)#0> |
| 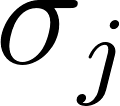<https://www.codecogs.com/eqnedit.php?latex=%5Csigma_j#0>  Rate per year of reactivation | 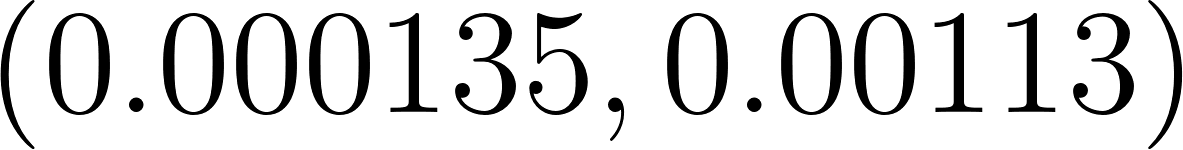<https://www.codecogs.com/eqnedit.php?latex=(0.000135%2C%200.00113)#0> | Retain if value for children is **less** than value for adults | Sample 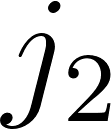<https://www.codecogs.com/eqnedit.php?latex=j_2#0> from 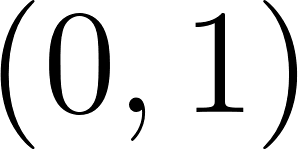<https://www.codecogs.com/eqnedit.php?latex=(0%2C1)#0> | Sample 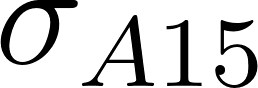<https://www.codecogs.com/eqnedit.php?latex=%5Csigma_%7BA15%7D#0> from  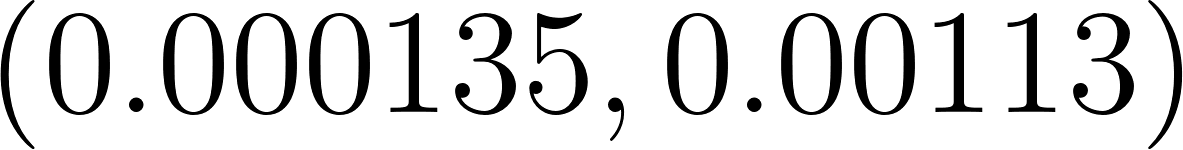<https://www.codecogs.com/eqnedit.php?latex=(0.000135%2C%200.00113)#0> | 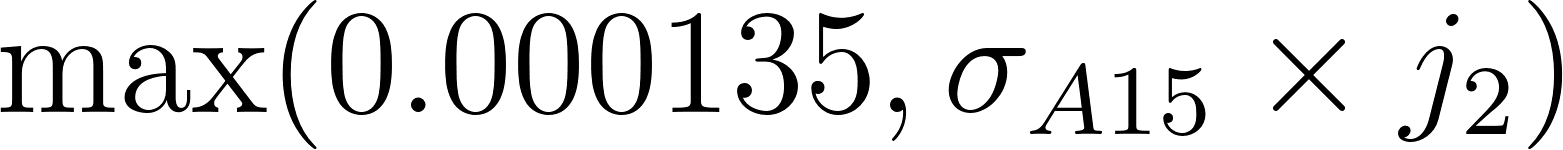<https://www.codecogs.com/eqnedit.php?latex=%5Cmax(0.000135%2C%20%5Csigma_%7BA15%7D%20%5Ctimes%20j_2)#0> |
| 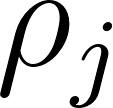<https://www.codecogs.com/eqnedit.php?latex=%5Crho_j#0>  Rate per year of relapse | 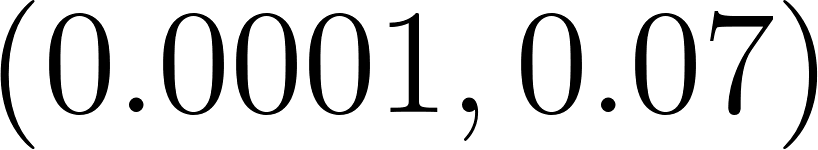<https://www.codecogs.com/eqnedit.php?latex=(0.0001%2C%200.07)#0> | Retain if value for children is **less** than value for adults | Sample 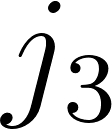<https://www.codecogs.com/eqnedit.php?latex=j_3#0> from 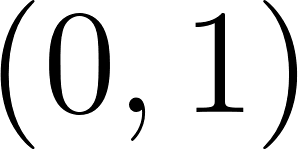<https://www.codecogs.com/eqnedit.php?latex=(0%2C1)#0> | Sample 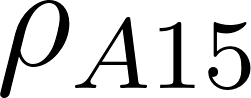<https://www.codecogs.com/eqnedit.php?latex=%5Crho_%7BA15%7D#0> from  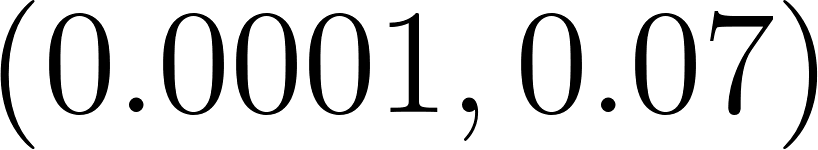<https://www.codecogs.com/eqnedit.php?latex=(0.0001%2C%200.07)#0> | 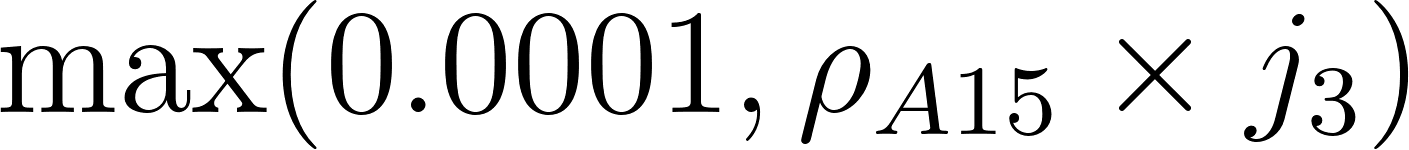<https://www.codecogs.com/eqnedit.php?latex=%5Cmax(0.0001%2C%20%5Crho_%7BA15%7D%20%5Ctimes%20j_3)#0> |
| 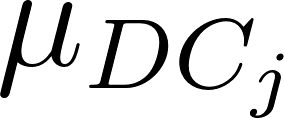<https://www.codecogs.com/eqnedit.php?latex=%5Cmu_%7BDC_j%7D#0>  Symptomatic TB mortality rate per year | 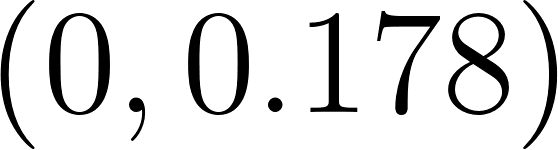<https://www.codecogs.com/eqnedit.php?latex=(0%2C%200.178)#0> | Retain if value for children is **greater** than value for adults | Sample 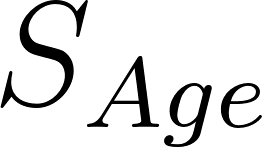<https://www.codecogs.com/eqnedit.php?latex=S_%7BAge%7D#0> from 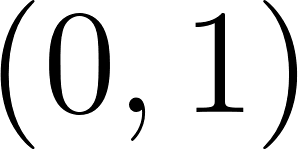<https://www.codecogs.com/eqnedit.php?latex=(0%2C1)#0> | 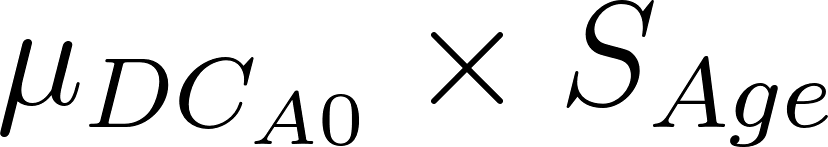<https://www.codecogs.com/eqnedit.php?latex=%5Cmu_%7BDC_%7BA0%7D%7D%20%5Ctimes%20S_%7BAge%7D#0> | Sample 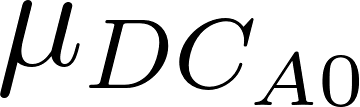<https://www.codecogs.com/eqnedit.php?latex=%5Cmu_%7BDC_%7BA0%7D%7D#0> from  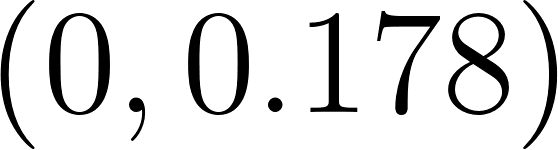<https://www.codecogs.com/eqnedit.php?latex=(0%2C%200.178)#0> |
| 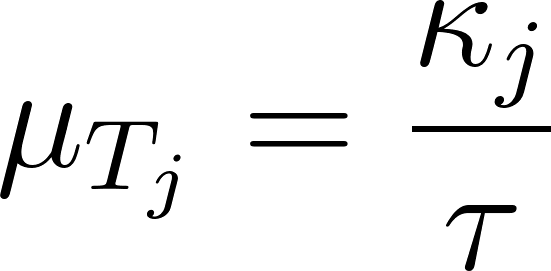<https://www.codecogs.com/eqnedit.php?latex=%5Cmu_%7BT_j%7D%20%3D%20%5Cfrac%7B%5Ckappa_j%7D%7B%5Ctau%7D#0>  On-treatment mortality rate per year | 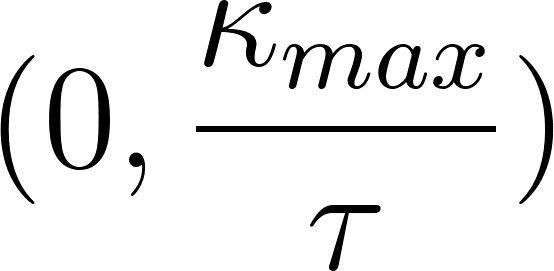<https://www.codecogs.com/eqnedit.php?latex=(0%2C%20%5Cfrac%7B%5Ckappa_%7Bmax%7D%7D%7B%5Ctau%7D)#0> | Retain if value for children is **greater** than value for adults | Sample 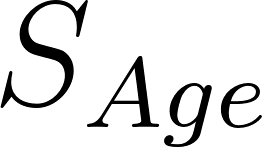<https://www.codecogs.com/eqnedit.php?latex=S_%7BAge%7D#0> from 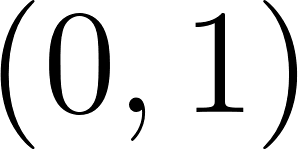<https://www.codecogs.com/eqnedit.php?latex=(0%2C1)#0> | 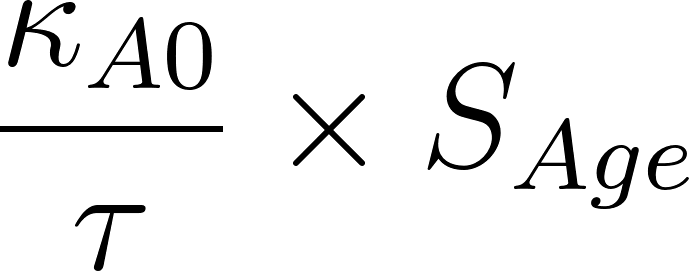<https://www.codecogs.com/eqnedit.php?latex=%20%5Cfrac%7B%5Ckappa_%7BA0%7D%7D%7B%5Ctau%7D%5Ctimes%20S_%7BAge%7D#0> | Sample 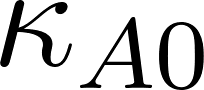<https://www.codecogs.com/eqnedit.php?latex=%5Ckappa_%7BA0%7D#0> from  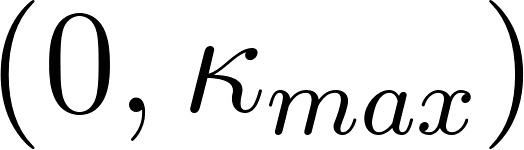<https://www.codecogs.com/eqnedit.php?latex=(0%2C%20%5Ckappa_%7Bmax%7D)#0> |

**TB treatment initiation and outcomes**

The approach for simulating TB treatment initiation and treatment outcomes is described in detail in Clark et al., 2023 and summarised here.^1^

The treatment initiation rate determines the transition from symptomatic TB to the on-treatment state. We assume that the availability of treatment started in 1960 and followed a sigmoidal increase to its current level in 2023. The rate of treatment initiation in 2023 was sampled during the calibration process. This is multiplied by the value of the sigmoidal curve (0–1) to give the value for each year. We also assume that the rate of treatment initiation is lower in children than adults. This was implemented via an additional scaling parameter sampled between 0–1 during calibration.

Our model has three possible outcomes while on treatment: death, treatment completion (transition to the R state), treatment non-completion (return to the sTB state). WHO reported data on outcomes is used to derive the rates of on-treatment mortality, treatment completion and treatment-non completion.^4^ First the fraction of deaths among children (>15 years old) starting treatment is sampled during calibration. This is scaled by a multiplier to give the fraction of deaths in adults. The fraction who do not die are then divided between completion and non-completion based on the ratio of completion to completion plus non-completion reported in the WHO data. Table S 3 below shows the calculations.

Table S 3	Calculating treatment outcome parameter values for adults and children

| **Parameter** | **Adults** | **Children** |
| --- | --- | --- |
| 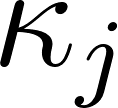<https://www.codecogs.com/eqnedit.php?latex=%20%5Ckappa_j#0>  On-treatment mortality fraction | 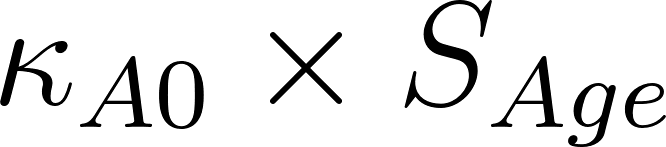<https://www.codecogs.com/eqnedit.php?latex=%5Ckappa_%7BA0%7D%5Ctimes%20S_%7BAge%7D#0> | Sample 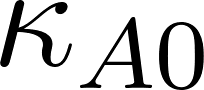<https://www.codecogs.com/eqnedit.php?latex=%5Ckappa_%7BA0%7D#0> from  0 to 2 x Average mortality on-treatment |
| 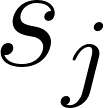<https://www.codecogs.com/eqnedit.php?latex=%20s_j#0>  On-treatment success fraction | 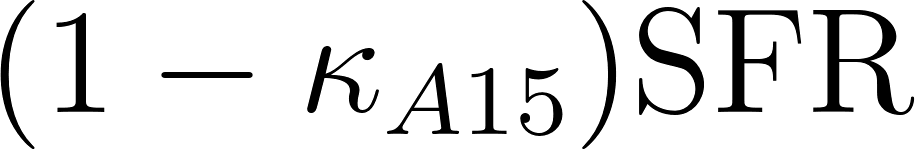<https://www.codecogs.com/eqnedit.php?latex=(1-%20%5Ckappa_%7BA15%7D)%20%5Ctext%7BSFR%7D#0> | 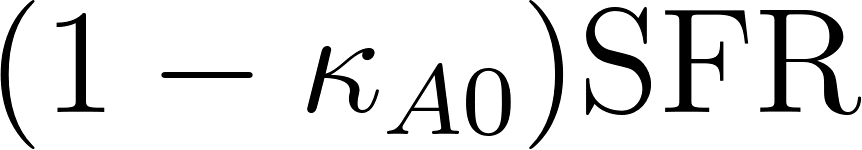<https://www.codecogs.com/eqnedit.php?latex=(1-%20%5Ckappa_%7BA0%7D)%20%5Ctext%7BSFR%7D#0> |
| 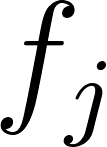<https://www.codecogs.com/eqnedit.php?latex=f_j#0>  On-treatment failure fraction | 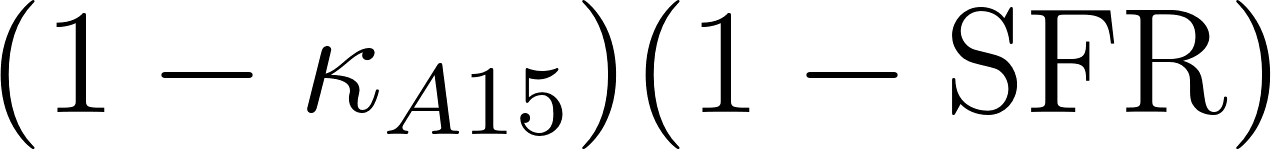<https://www.codecogs.com/eqnedit.php?latex=(1-%20%5Ckappa_%7BA15%7D)%20(1-%5Ctext%7BSFR%7D)#0> | 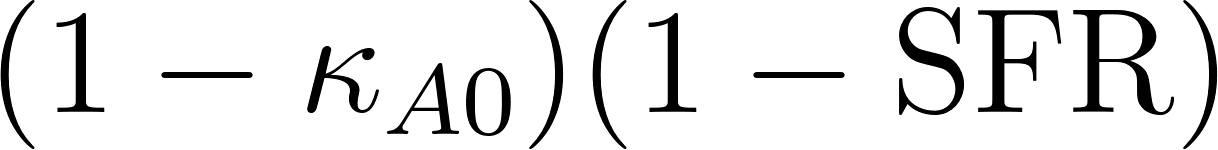<https://www.codecogs.com/eqnedit.php?latex=(1-%20%5Ckappa_%7BA0%7D)%20(1-%5Ctext%7BSFR%7D)#0> |

Then divide each of the parameters by
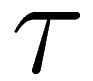
<http://www.sciweavers.org/tex2img.php?bc=Transparent&fc=Black&im=jpg&fs=100&ff=modern&edit=0&eq=%5Ctau#0> (the treatment duration parameter) to get the on-treatment mortality rate per year, on-treatment success rate per year, and on-treatment failure rate per year.

### 2. Model simulation and calibration

The model consists of a system of ordinary differential equations which define the derivatives with respect to time of the state variables. The transitions between state variables, together with the parameters and other model inputs, are specified in an xml file using a standardised schema. The equations are then generated and solved using a matrix multiplication approach. This is implemented in the R programming language.

The model is used to simulate the progression of the TB and HIV epidemics from 1900 to 2050. The model is initialised by distributing the population between the TB disease states using a parameter representing the proportion of the population uninfected at the start of the simulation and initially simulated without new TB vaccines to establish the baseline no-new-vaccine scenario. For each year of the baseline simulation the birth rate and age-specific mortality rates are adjusted to match the UN population estimates and projections. This allows us to account for TB and HIV mortality, which is simulated independently in our model but implicitly included in the UN estimates and projections. The baseline, with the adjusted birth and mortality rates, is then used to simulate different vaccination scenarios.

To establish the no-new-vaccine baseline the model was calibrated to a set of TB (and HIV) calibration targets for each country (Table S 4 and Table S 5). Calibration was carried out using history matching with emulation, a method that allows us to explore high-dimensional parameter spaces efficiently and robustly. History matching progresses as a series of iterations, called waves, where implausible areas of the parameter space, i.e., areas that are unable to give a match between the model output (e.g., the predicted incidence rate by the model) and the empirical data (e.g., the incidence rate calibration target from the WHO data), are found and discarded. In order to identify implausible parameter sets, emulators, which are statistical approximations of model outputs that are built using a modest number of model runs, are used (in this example we used 400 model runs, 200 for training and 200 for validation).

Emulators provide an estimate of the value of the model at any parameter set of interest, with the advantage that they are orders of magnitude faster than the model. History matching with emulation, implemented through the *hmer* package in R,^21–23^ considerably reduced the size of the parameter space to investigate. Rejection sampling was then performed on the reduced space to identify at least 200 parameter sets that matched all targets.

For two countries unable to find at least 200 fully fitted parameter sets using history matching with emulation, they were subsequently assessed using an Approximate Bayesian Computation Markov Chain Monte Carlo method (ABC-MCMC). ABC-MCMC was conducted using the *easyABC* package in R, modified by the Sebastian Funk, Gwenan Knight, and the TB Modelling Group at LSHTM for adaptive sampling and to accept seeded parameter values.^24,25^ We used parameter sets with the maximum number of targets fitted during history matching with emulation as starting seeds, with the ABC-MCMC algorithm continuously adapting using the last 1000 points, a burn in of 1000 samples, and the noise factor set to 0.0001. Once we had obtained 200 parameter sets that produced output consistent with the calibration targets, we used those parameter sets with the mechanistic model to simulate the future trajectories of TB.

##### **Country-specific calibration targets**

The model was calibrated separately for each of the 79 countries to eight calibration targets, with countries classified as having a high HIV/TB burden calibrated to at least three additional TB-HIV or HIV specific calibration targets. The seven TB calibration targets that varied by country are listed in Table S 4 below, and the additional calibration targets for countries classified as having a high HIV/TB burden are included in Table S 5. The eighth calibration target included for all 79 countries was the proportion of infectious TB that was asymptomatic (0.504 [0.361, 0.797] from Frascella, et al.^26^).

Table S 4	TB specific calibration targets for the 79 included countries.

| **Country Code** | **TB incidence rate per 100,000 population in 2023**^4,27^ | | | **TB case notification rate per 100,000 population in 2023**^4,27^ | | | **TB mortality rate per 100,000 population in 2023 (all ages)**^4^ |
| --- | --- | --- | --- | --- | --- | --- | --- |
|  | **All ages** | **Ages 0–14** | **Ages 15–99** | **All ages** | **Ages 0–14** | **Ages 15–99** |  |
| AFG | 180 (112, 263) | 78.2 (40.2, 122.9) | 252.4 (126.2, 382.8) | 119.0 (95.2, 142.8) | 60.8 (48.64, 72.96) | 162.5 (130.0, 195.0) | 24 (15, 37) |
| AGO | 339 (214, 520) | 117.1 (56.1, 178.7) | 527.7 (251.3, 809.2) | 177.0 (141.6, 212.4) | 46.6 (37.28, 55.92) | 281.6 (225.28, 337.92) | 61 (38, 90) |
| ARG | 35 (30, 41) | 12.4 (10.5, 14.4) | 42.7 (34.1, 48.3) | 31.0 (24.8, 37.2) | 10.5 (8.4, 12.6) | 36.1 (28.88, 43.32) | 1.8 (1.6, 2) |
| AZE | 72 (48, 107) | 17.8 (10.4, 25.7) | 87.8 (50.2, 125.4) | 38.0 (30.4, 45.6) | 5.8 (4.64, 6.96) | 48 (38.4, 57.6) | 5.2 (4.6, 5.8) |
| BDI | 94 (58, 138) | 20.2 (10.6, 30.3) | 168.5 (85.7, 238.7) | 58.0 (46.4, 69.6) | 5.8 (4.64, 6.96) | 107.9 (86.32, 129.48) | 18 (11, 27) |
| BEN | 51 (31, 75) | 12.0 (6.3, 17.4) | 83.3 (43.6, 121.8) | 31.0 (24.8, 37.2) | 3.2 (2.56, 3.84) | 52.9 (42.32, 63.48) | 11 (6.8, 15) |
| BFA | 43 (27, 63) | 9.7 (5.2, 14) | 68.7 (37.0, 100.3) | 38.0 (30.4, 45.6) | 3 (2.4, 3.6) | 64.5 (51.6, 77.4) | 3.6 (2.1, 5.5) |
| BGD | 221 (161, 291) | 76.8 (51.9, 103.9) | 270 (183.1, 356.8) | 177 (141.6, 212.4) | 30 (24, 36) | 226.6 (181.28, 271.92) | 26 (16, 39) |
| BOL | 105 (67, 153) | 22.8 (12.7, 31.8) | 140.7 (78.6, 199.4) | 76.0 (60.8, 91.2) | 7.6 (6.08, 9.12) | 106 (84.8, 127.2) | 11 (8, 15) |
| BRA | 49 (41, 56) | 8.1 (6.7, 9.2) | 58.0 (48.7, 67.3) | 44.0 (35.2, 52.8) | 7.1 (5.68, 8.52) | 51.7 (41.36, 62.04) | 6.1 (5.3, 7.1) |
| BTN | 164 (126, 208) | 19.8 (14.6, 24.5) | 211.9 (153.2, 260.8) | 107.0 (85.6, 128.4) | 12.8 (10.24, 15.36) | 133.3 (106.64, 159.96) | 28 (19, 39) |
| CHN | 52 (44, 61) | 14.0 (11.5, 16.5) | 59.8 (49.7, 69.7) | 40.0 (32.0, 48.0) | 2.7 (2.16, 3.24) | 47.2 (37.76, 56.64) | 1.9 (1.7, 2.1) |
| COD | 316 (204, 453) | 102.5 (55.5, 149.5) | 531.7 (290.0, 773.4) | 244 (195.2, 292.8) | 73.9 (59.12, 88.68) | 415.1 (332.08, 498.12) | 41 (26, 59) |
| COG | 368 (228, 539) | 105.4 (56.8, 158.1) | 560.0 (308.0, 840.0) | 232 (185.6, 278.4) | 45 (36, 54) | 371.2 (296.96, 445.44) | 84 (54, 119) |
| COL | 46 (33, 62) | 8.9 (6.1, 11.8) | 56.2 (39.1, 73.3) | 38 (30.4, 45.6) | 5.70 (4.56, 6.84) | 47.3 (37.84, 56.76) | 4.4 (3.6, 5) |
| CPV | 47 (36, 60) | 5.8 (3.9, 7.1) | 54.4 (40.8, 68) | 38 (30.4, 45.6) | 4.50 (3.60, 5.40) | 43.1 (34.48, 51.72) | 5.8 (4, 7.9) |
| DOM | 42 (31, 54) | 4.9 (3.6, 6.6) | 55.8 (40.1, 71.6) | 39 (31.2, 46.8) | 4.70 (3.76, 5.64) | 51.4 (41.12, 61.68) | 4.1 (2.4, 6.1) |
| ECU | 58 (44, 73) | 15.8 (11.7, 20.2) | 71.2 (52.7, 89) | 46 (36.8, 55.2) | 9.3 (7.44, 11.16) | 58.4 (46.72, 70.08) | 4.7 (3.9, 5.4) |
| EGY | 9.2 (7.8 to 11) | 1.7 (1.4 to 2) | 13.2 (11 to 16) | 8.1 (6.48 to 9.72) | 1.5 (1.2 to 1.8) | 11.6 (9.28 to 13.92) | 0.46 (0.41 to 0.52) |
| ERI | 65 (23 to 127) | 32.5 (0.5 to 64.3) | 79.3 (1.1 to 154.2) | 64 (51.2 to 76.8) | 32.2 (25.76 to 38.64) | 77.3 (61.84 to 92.76) | 2.3 (0.41 to 5.9) |
| ETH | 146 (98 to 203) | 46.7 (28.4 to 66.9) | 216.8 (129.6 to 305.4) | 105 (84 to 126) | 28.2 (22.56 to 33.84) | 159.9 (127.92 to 191.88) | 23 (15 to 32) |
| FJI | 66 (51, 84) | 14.7 (10.9, 18.8) | 85.4 (62.9, 109.4) | 42 (33.6, 50.4) | 9.4 (7.52, 11.28) | 54.8 (43.84, 65.76) | 5.3 (4.9, 5.7) |
| GAB | 505 (318, 733) | 137.4 (75.6, 206.1) | 714.7 (402.8, 1039.5) | 268 (214.4, 321.6) | 31.8 (25.44, 38.16) | 413.9 (331.12, 496.68) | 139 (84, 208) |
| GHA | 129 (57, 231) | 48.3 (11.3, 88.5) | 177.8 (40.2, 313.6) | 56 (44.8, 67.2) | 6.6 (5.28, 7.92) | 84.3 (67.44, 101.16) | 36 (18, 61) |
| GIN | 175 (111, 252) | 39.6 (22.4, 55.1) | 279.8 (158.2, 401.5) | 139 (111.2, 166.8) | 20.8 (16.64, 24.96) | 229.3 (183.44, 275.16) | 23 (16, 32) |
| GMB | 142 (101, 189) | 24.7 (16.2, 32.4) | 223.5 (153.3, 300.2) | 103 (82.4, 123.6) | 12.6 (10.08, 15.12) | 167.2 (133.76, 200.64) | 22 (16, 30) |
| GNQ | 274 (175, 396) | 53.9 (30.8, 77) | 449.8 (258.4, 650.7) | 197 (157.6, 236.4) | 31.9 (25.52, 38.28) | 329.2 (263.36, 395.04) | 63 (45, 85) |
| GTM | 33 (25, 41) | 15.4 (10.9, 19.1) | 41 (29.5, 53.3) | 26 (20.8, 31.2) | 12 (9.6, 14.4) | 33 (26.4, 39.6) | 3 (2.6, 3.4) |
| IDN | 387 (354, 432) | 335 (293.1, 375.4) | 413.8 (363.1, 464.9) | 286 (228.8, 343.2) | 197.2 (157.76, 236.64) | 322.6 (258.08, 387.12) | 47 (40, 53) |
| IND | 195 (164, 228) | 87.1 (71.1, 102.8) | 233.8 (191.6, 277) | 166 (132.8, 199.2) | 37.9 (30.32, 45.48) | 211.1 (168.88, 253.32) | 22 (16, 30) |
| IRN | 11 (7.9, 14) | 2.2 (1.5, 2.8) | 13.8 (9.6, 17.6) | 8.2 (6.56, 9.84) | 1.0 (0.8, 1.2) | 10.6 (8.48, 12.72) | 1.1 (1.1, 1.2) |
| IRQ | 21 (17, 25) | 6.5 (5, 7.7) | 29.5 (23.8, 35.6) | 15.0 (12.0, 18.0) | 3.1 (2.48, 3.72) | 22.8 (18.24, 27.36) | 1.6 (1.4, 1.8) |
| JOR | 3.4 (2.3, 4.7) | 0.9 (0.5, 1.2) | 4.7 (2.9, 6.3) | 2.2 (1.76, 2.64) | 0.6 (0.48, 0.72) | 3 (2.4, 3.6) | 0.09 (0.07, 0.1) |
| KAZ | 70 (43, 120) | 13.6 (5.7, 20.7) | 94.8 (40.8, 153.2) | 47.0 (37.6, 56.4) | 5.4 (4.32, 6.48) | 67.1 (53.68, 80.52) | 3.2 (2.2, 4.2) |
| KGZ | 112 (88, 134) | 31.8 (24.4, 38.7) | 164.2 (125.5, 200.7) | 59.0 (47.2, 70.8) | 11.8 (9.44, 14.16) | 89.2 (71.36, 107.04) | 5.2 (4.8, 5.9) |
| KHM | 335 (212, 494) | 247.5 (119.6, 371.3) | 383.1 (191.6, 583) | 185.0 (148.0, 222.0) | 147.5 (118, 177) | 208.2 (166.56, 249.84) | 25 (16, 35) |
| LAO | 132 (81, 197) | 36.4 (19.1, 52) | 176.4 (92.9, 265.5) | 119.0 (95.2, 142.8) | 4.5 (3.6, 5.4) | 171.1 (136.88, 205.32) | 9.9 (5.3, 16) |
| LBR | 308 (197, 444) | 115.8 (64.9, 171.5) | 437.3 (240.5, 655.9) | 134.0 (107.2, 160.8) | 37.1 (29.68, 44.52) | 204.9 (163.92, 245.88) | 85 (54, 123) |
| LBY | 59 (36, 87) | 16.2 (8.9, 23.5) | 81.1 (42.6, 117.6) | 32.0 (25.6, 38.4) | 5.7 (4.56, 6.84) | 44.8 (35.84, 53.76) | 13 (7.7, 20) |
| LKA | 62 (45, 82) | 14.6 (9.9, 19) | 82.7 (55, 106.4) | 40.0 (32.0, 48.0) | 4.3 (3.44, 5.16) | 53.5 (42.8, 64.2) | 3.6 (3.2, 3.9) |
| LSO | 664 (335, 985) | 178.1 (85.2, 279.9) | 914.2 (417.9, 1371.3) | 281.0 (224.8, 337.2) | 46.7 (37.36, 56.04) | 400.2 (320.16, 480.24) | 229 (154, 298) |
| MAR | 92 (77, 108) | 25.1 (21.1, 30.1) | 115.5 (93.9, 137.2) | 86.0 (68.8, 103.2) | 23.7 (18.96, 28.44) | 108.6 (86.88, 130.32) | 5.1 (2.5, 8.7) |
| MDA | 76 (64, 88) | 24.3 (21.3, 28.9) | 77.5 (63.4, 88.0) | 71.0 (56.8, 85.2) | 23.3 (18.64, 27.96) | 70.8 (56.64, 84.96) | 6.7 (6, 7.2) |
| MDG | 233 (149, 336) | 74.6 (40.3, 111.5) | 349.6 (191.2, 508.0) | 149.0 (119.2, 178.8) | 33.8 (27.04, 40.56) | 232.1 (185.68, 278.52) | 42 (25, 62) |
| MEX | 29 (20, 42) | 5.8 (3.2, 8.1) | 36.1 (21.7, 51.6) | 21.0 (16.8, 25.2) | 2.5 (2, 3) | 27.1 (21.68, 32.52) | 3.5 (2.9, 4.5) |
| MMR | 558 (328, 824) | 330.1 (165, 502.6) | 628.5 (306.9, 950.1) | 239.0 (191.2, 286.8) | 171.9 (137.52, 206.28) | 258.7 (206.96, 310.44) | 90 (62, 125) |
| MNG | 491 (280, 778) | 108.1 (50.5, 171.2) | 691.6 (306.9, 1037.4) | 87.0 (69.6, 104.4) | 15 (12, 18) | 122.4 (97.92, 146.88) | 12 (11, 15) |
| MOZ | 361 (220, 537) | 117.1 (56.5, 172.2) | 555.4 (275.1, 835.7) | 346.0 (276.8, 415.2) | 90 (72, 108) | 546.1 (436.88, 655.32) | 23 (12, 37) |
| MRT | 74 (44, 112) | 17.6 (8.6, 26.2) | 120.9 (60.4, 181.3) | 49.0 (39.2, 58.8) | 6.1 (4.88, 7.32) | 82.2 (65.76, 98.64) | 13 (7.4, 20) |
| MWI | 119 (55, 206) | 30.9 (8.9, 52.6) | 184.6 (54.5, 318.8) | 89.0 (71.2, 106.8) | 15.3 (12.24, 18.36) | 146.4 (117.12, 175.68) | 23 (14, 34) |
| MYS | 122 (87, 155) | 41.6 (28.6, 54.6) | 151.3 (105.9, 196.7) | 74.0 (59.2, 88.8) | 21.8 (17.44, 26.16) | 92.6 (74.08, 111.12) | 8.9 (5.4, 13) |
| NAM | 468 (221, 622) | 181.4 (92.8, 266.7) | 728.4 (382.4, 1092.6) | 312.0 (249.6, 374.4) | 99.4 (79.52, 119.28) | 504.9 (403.92, 605.88) | 98 (52, 132) |
| NER | 74 (45, 111) | 10 (5.2, 14.6) | 131.7 (70.3, 197.6) | 59.0 (47.2, 70.8) | 5.5 (4.4, 6.6) | 108 (86.4, 129.6) | 8.5 (4.9, 13) |
| NGA | 219 (143, 311) | 62.2 (34.8, 88.5) | 349.3 (198.8, 499.8) | 161.0 (128.8, 193.2) | 38 (30.4, 45.6) | 262.1 (209.68, 314.52) | 31 (20, 45) |
| NPL | 229 (126, 355) | 59 (27.2, 91.8) | 287.6 (132.4, 442.8) | 124.0 (99.2, 148.8) | 33.8 (27.04, 40.56) | 154.8 (123.84, 185.76) | 55 (29, 86) |
| PAK | 277 (188, 368) | 108.6 (68.1, 149) | 391.3 (246.1, 536.5) | 192.0 (153.6, 230.4) | 77.5 (62.0, 93.0) | 269.6 (215.68, 323.52) | 20 (16, 24) |
| PER | 173 (114, 250) | 45.3 (26, 64.5) | 217 (126.2, 307.7) | 94.0 (75.2, 112.8) | 15.4 (12.32, 18.48) | 119.6 (95.68, 143.52) | 16 (13, 22) |
| PHL | 643 (296, 1120) | 170.8 (54.1, 290.3) | 835 (258.3, 1414.3) | 501.0 (400.8, 601.2) | 147.7 (118.16, 177.24) | 644.2 (515.36, 773.04) | 32 (22, 45) |
| PNG | 432 (348, 525) | 344 (255.1, 430) | 489 (355.6, 622.4) | 393.0 (314.4, 471.6) | 270.1 (216.08, 324.12) | 461 (368.8, 553.2) | 31 (18, 47) |
| PRY | 62 (53, 72) | 7.6 (6.6, 8.7) | 84.5 (72.1, 98.9) | 54.0 (43.2, 64.8) | 6.6 (5.28, 7.92) | 73.9 (59.12, 88.68) | 4.3 (3.7, 4.8) |
| SDN | 50 (31, 75) | 20.6 (10.3, 30.4) | 74.7 (39.2, 113.9) | 27.0 (21.6, 32.4) | 5.9 (4.72, 7.08) | 39.7 (31.76, 47.64) | 11 (6.5, 17) |
| SEN | 110 (76, 150) | 35.9 (22.1, 49.7) | 165.2 (106.9, 233.2) | 92.0 (73.6, 110.4) | 11.0 (8.8, 13.2) | 154.2 (123.36, 185.04) | 11 (6.8, 16) |
| SLV | 84 (65, 107) | 20.6 (15, 26.2) | 105.3 (77.9, 132.7) | 68.0 (54.4, 81.6) | 10.1 (8.08, 12.12) | 86.5 (69.2, 103.8) | 2.3 (2, 2.6) |
| SUR | 29 (22, 36) | 4.3 (3.7, 5.6) | 37.1 (28.4, 48) | 19.0 (15.2, 22.8) | 3.1 (2.48, 3.72) | 25.1 (20.08, 30.12) | 4.1 (3.3, 5) |
| SWZ | 350 (145, 507) | 122.4 (50.4, 194.4) | 481.5 (202.7, 760.3) | 187.0 (149.6, 224.4) | 47.5 (38, 57) | 266 (212.8, 319.2) | 77 (36, 114) |
| SYR | 17 (12, 22) | 4.5 (3.2, 5.9) | 23.5 (15.9, 30.5) | 14.0 (11.2, 16.8) | 3.8 (3.04, 4.56) | 19.9 (15.92, 23.88) | 0.14 (0.12, 0.15) |
| TCD | 139 (88, 202) | 38.7 (21.1, 56.2) | 253.7 (137.4, 359.4) | 82.0 (65.6, 98.4) | 11.3 (9.04, 13.56) | 156.5 (125.2, 187.8) | 30 (19, 43) |
| THA | 157 (114, 214) | 35.4 (23.6, 47.1) | 179.6 (120.3, 238.9) | 112.0 (89.6, 134.4) | 5.90 (4.72, 7.08) | 131.1 (104.88, 157.32) | 19 (15, 25) |
| TJK | 79 (60, 99) | 17 (12.6, 21.7) | 117.0 (85.8, 148.2) | 43.0 (34.4, 51.6) | 7.5 (6, 9) | 64.9 (51.92, 77.88) | 8.3 (7.4, 9.2) |
| TLS | 498 (322, 711) | 156.9 (88.1, 214.9) | 700.1 (395.2, 993.7) | 435.0 (348.0, 522.0) | 60.2 (48.16, 72.24) | 649 (519.2, 778.8) | 40 (22, 64) |
| TUN | 38 (29, 47) | 12.0 (8.8, 15.3) | 45.0 (33.2, 56.8) | 26.0 (20.8, 31.2) | 6.7 (5.36, 8.04) | 31.2 (24.96, 37.44) | 1.3 (0.93, 1.6) |
| UKR | 112 (66, 170) | 35.2 (17.6, 52.8) | 131.7 (69.1, 197.5) | 53.0 (42.4, 63.6) | 11.2 (8.96, 13.44) | 63.9 (51.12, 76.68) | 17.0 (12.0, 22.0) |
| VEN | 45 (34, 58) | 11.3 (8.0, 14.2) | 57.7 (39.9, 72.1) | 36.0 (28.8, 43.2) | 6.9 (5.52, 8.28) | 46.3 (37.04, 55.56) | 3.2 (2.6, 4.0) |
| VNM | 182 (113, 264) | 33.7 (19.1, 50.1) | 228.6 (129.3, 327.9) | 104.0 (83.2, 124.8) | 5.3 (4.24, 6.36) | 134.9 (107.92, 161.88) | 13.0 (8.2, 18.0) |
| VUT | 41 (31, 53) | 6.2 (4.6, 7.7) | 59.8 (44.8, 79.7) | 32.0 (25.6, 38.4) | 4.6 (3.68, 5.52) | 47.3 (37.84, 56.76) | 5.3 (3.5, 7.5) |
| YEM | 48 (41, 55) | 19.4 (16.4, 22.4) | 77.4 (67.7, 91.9) | 28.0 (22.4, 33.6) | 9.6 (7.68, 11.52) | 47.0 (37.6, 56.4) | 9.5 (6.8, 13.0) |
| ZAF | 427 (265, 626) | 134.3 (75.9, 198.6) | 574.2 (309.2, 837.0) | 335.0 (268, 402) | 86.4 (69.12, 103.68) | 458 (366.4, 549.6) | 88.0 (48.0, 139.0) |
| ZMB | 283 (173, 420) | 102.8 (50.8, 150.2) | 429.8 (206.3, 644.7) | 262.0 (209.6, 314.4) | 71.8 (57.44, 86.16) | 413.7 (330.96, 496.44) | 24.0 (15.0, 35.0) |
| ZWE | 211 (135, 284) | 65.9 (40.4, 92.9) | 305.7 (183.4, 428.0) | 118.0 (94.4, 141.6) | 17 (13.6, 20.4) | 184.2 (147.36, 221.04) | 48.0 (34.0, 63.0) |

Table S 5	Calibration targets for the 10 countries classified as having a high HIV/TB burden.

| **Country Code** | **TB incidence rate in PLHIV per 100,000 population in 2023 for all ages**^4^ | **TB mortality rate in PLHIV per 100,000 population in 2023 for all ages**^4^ | **HIV prevalence in 2021 for all ages (%)**^28^ | **ART coverage in 2021 for all ages  (%)**^28^ |
| --- | --- | --- | --- | --- |
| COG | 112 (55, 189) | 38 (19, 64) | 2.2 (1.6, 3.3) | 23 (16, 35) |
| GAB | 150 (61, 278) | 62 (25, 115) | 1.9 (1.7, 2.4) | 54 (46, 69) |
| GNQ | 173 (110, 249) | 49 (32, 70) | 3.9 (3.8, 4.2) | 41 (40, 44) |
| LSO | 362 (199, 574) | 161 (102, 234) | 12.5 (11.7, 13.4) | 81 (77, 87) |
| MWI | 56 (26, 98) | 15 (7.7, 24) | 4.8 (4.5, 5.3) | 91 (85, 99) |
| NAM | 136 (80, 207) | 44 (24, 70) | 8.5 (7.7, 9.3) | 91 (85, 100) |
| SWZ | 188 (91, 318) | 53 (25, 93) | 18.2 (17.4, 19.1) | 91 (85, 97) |
| ZAF | 230 (143, 337) | 49 (14, 104) | 12.5 (11.6, 13.6) | 74 (68, 80) |
| ZMB | 91 (55, 135) | 13 (7, 20) | 6.4 (6.4, 6.9) | 90 (86, 97) |
| ZWE | 127 (80, 184) | 34 (22, 48) | 7.9 (7.3, 8.5) | 91 (84, 100) |

### 3. Vaccine delivery

We modelled eight vaccine delivery scenarios with specific characteristics as listed in Table 1 in the main text. We assumed that the vaccine would impact progression to infectious disease by multiplying all rates of flow into the asymptomatic TB compartment by (1-pV), where pV is the estimate of vaccine efficacy. In our modelling, we assumed that there was no pre-vaccination infection testing. Therefore, for scenarios where the vaccine is only effective when delivered to uninfected individuals at the time of vaccination, we assumed that both uninfected and infected individuals would receive the vaccine, and only the uninfected individuals would receive protection. Our model structure allows for counting and tracking individuals who received the vaccine but do not receive any protection from it.

### Vaccine efficacy estimates

Reported vaccine efficacy estimates from the VPM1002 POD trial are included in Table 1 in the main text.^29^ To represent the underlying uncertainty, for each of the 200 model simulations of a vaccine scenario, we sampled from the corresponding distribution of the range of the vaccine efficacy as below:

1. Using the reported hazard ratio, 95% confidence intervals, and total number of events for each vaccine efficacy estimate, we simulated the distribution of the log hazard ratio.
2. We sampled from the distribution of the log hazard ratio.
3. We translated those sampled estimates into vaccine efficacy values which we used for the simulation.

Distributions below are for the *Main analysis*, *Young* and *Uninfected* scenarios, but the same procedure was used for all analyses.

***Main analysis* scenario**

The hazard ratio from the trial was 0.831 (0.609 to 1.133), leading to a vaccine efficacy estimate of 0.169 (-0.133 to 0.391).^29^ The dashed blue line in Figure S 3 represents the true underlying distribution being sampled and the solid black line represents the sampled distribution. The solid vertical line represents the median estimates from the trial and simulation, and the dashed vertical lines represent the 95% confidence limits from the trial and simulation.

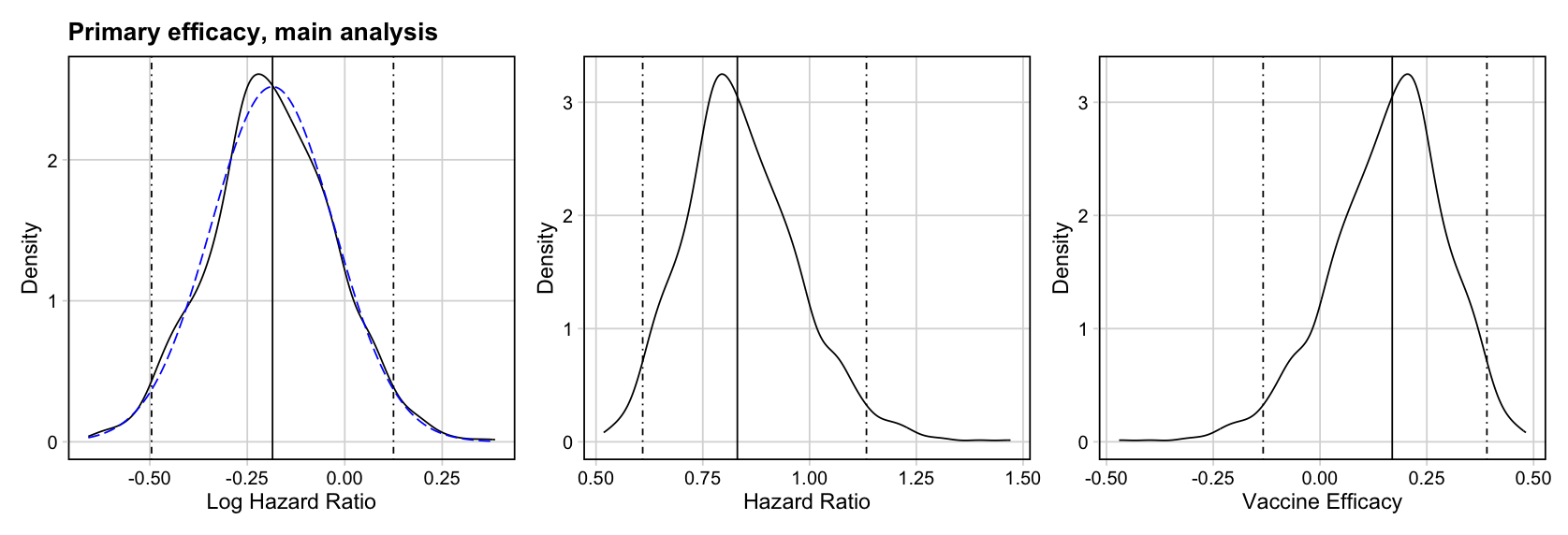

Figure S 3	The simulated and true log hazard ratio distribution for the *Main analysis* scenario

***Young* scenario**

The hazard ratio from the trial was 0.728 (0.461 to 1.148), leading to a vaccine efficacy estimate of 0.272 (-0.148 to 0.539).^29^ The dashed blue line in Figure S 4 represents the true underlying distribution being sampled and the solid black line represents the sampled distribution. The solid vertical line represents the median estimates from the trial and simulation, and the dashed vertical lines represent the 95% confidence limits from the trial and simulation.

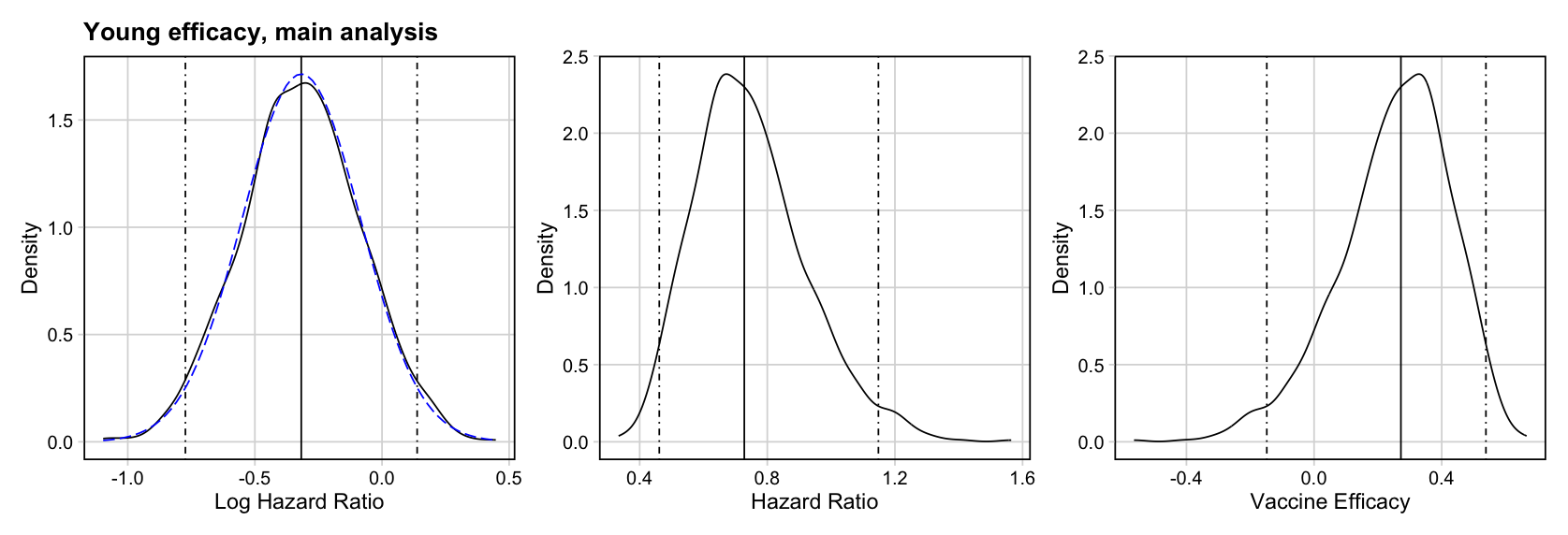

Figure S 4	The simulated and true log hazard ratio distribution for the *Young* scenario

***Uninfected* scenario**

The hazard ratio from the trial was 0.610 (0.362 to 1.027), leading to a vaccine efficacy estimate of 0.390 (-0.270 to 0.638).^29^ The dashed blue line in Figure S 5 represents the true underlying distribution being sampled and the solid black line represents the sampled distribution. The solid vertical line represents the median estimates from the trial and simulation, and the dashed vertical lines represent the 95% confidence limits from the trial and simulation.

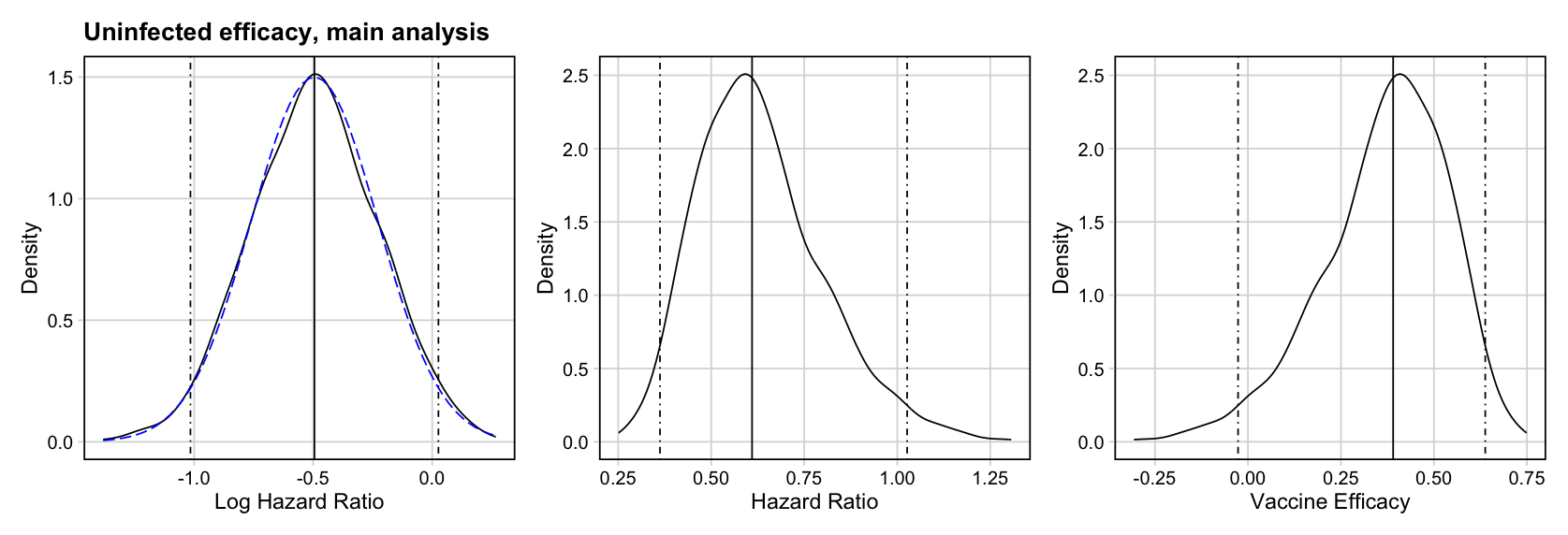

Figure S 5	The simulated and true log hazard ratio distribution for the *Uninfected* scenario

### Country-specific vaccine introduction years

The country-specific introduction years used in the analysis were as in the WHO investment case for new TB vaccines, shifted forward for the earliest delivery to occur in 2027.^30^ The specific introduction year for each country is listed in Table S6 below.

Table S 6	Country specific introduction years for the 79 LMICs included in the analysis

| **Country Code** | **Country-specific introduction year** |
| --- | --- |
| AFG | 2030 |
| AGO | 2031 |
| ARG | 2030 |
| AZE | 2027 |
| BDI | 2043 |
| BEN | 2036 |
| BFA | 2038 |
| BGD | 2034 |
| BOL | 2036 |
| BRA | 2029 |
| BTN | 2033 |
| CHN | 2028 |
| COD | 2029 |
| COG | 2034 |
| COL | 2029 |
| CPV | 2042 |
| DOM | 2030 |
| ECU | 2032 |
| EGY | 2032 |
| ERI | 2046 |
| ETH | 2029 |
| FJI | 2030 |
| GAB | 2037 |
| GHA | 2039 |
| GIN | 2032 |
| GMB | 2038 |
| GNQ | 2041 |
| GTM | 2035 |
| IDN | 2033 |
| IND | 2032 |
| IRN | 2030 |
| IRQ | 2032 |
| JOR | 2036 |
| KAZ | 2027 |
| KGZ | 2043 |
| KHM | 2035 |
| LAO | 2034 |
| LBR | 2036 |
| LBY | 2034 |
| LKA | 2027 |
| LSO | 2038 |
| MAR | 2028 |
| MDA | 2033 |
| MDG | 2030 |
| MEX | 2028 |
| MMR | 2030 |
| MNG | 2031 |
| MOZ | 2031 |
| MRT | 2041 |
| MWI | 2037 |
| MYS | 2027 |
| NAM | 2029 |
| NER | 2035 |
| NGA | 2029 |
| NPL | 2035 |
| PAK | 2030 |
| PER | 2028 |
| PHL | 2029 |
| PNG | 2031 |
| PRY | 2034 |
| SDN | 2035 |
| SEN | 2037 |
| SLV | 2038 |
| SUR | 2039 |
| SWZ | 2035 |
| SYR | 2035 |
| TCD | 2032 |
| THA | 2030 |
| TJK | 2044 |
| TLS | 2030 |
| TUN | 2035 |
| UKR | 2032 |
| VEN | 2034 |
| VNM | 2037 |
| VUT | 2041 |
| YEM | 2035 |
| ZAF | 2028 |
| ZMB | 2033 |
| ZWE | 2031 |

**Supplementary Results**

**Table S 7	Vaccine efficacy estimates and characteristics from the VPM1002 POD trial^29^**

| **Analysis (name assigned by this paper’s authors, not trialists)** | **Outcome** | **Cohort** | **TST Status** | **Age Group** | **Efficacy  (95% confidence interval)** |
| --- | --- | --- | --- | --- | --- |
| Main analysis | Microbiologically confirmed EPTB + PTB | mITT | Any | Ages 6+ | 16.9%  (-13.3, 39.1) |
| Per protocol | Microbiologically confirmed EPTB + PTB | Per protocol | Any | Ages 6+ | 21.4%  (-8.9, 43.2) |
| Prevents PTB-only | Microbiologically confirmed PTB | mITT | Any | Ages 6+ | 13.6%  (-21.1, 38.3) |
| Young | Microbiologically confirmed EPTB + PTB | mITT | Any | Age 6 to 18 | 27.2%  (-14.8, 53.9) |
| Uninfected | Microbiologically confirmed EPTB + PTB | mITT | Negative at baseline | Ages 6+ | 39.0%  (-2.7, 63.8) |

*Abbreviations: EPTB = extrapulmonary tuberculosis; mITT = modified intention to treat; PTB = pulmonary tuberculosis, TST = tuberculin skin test.*

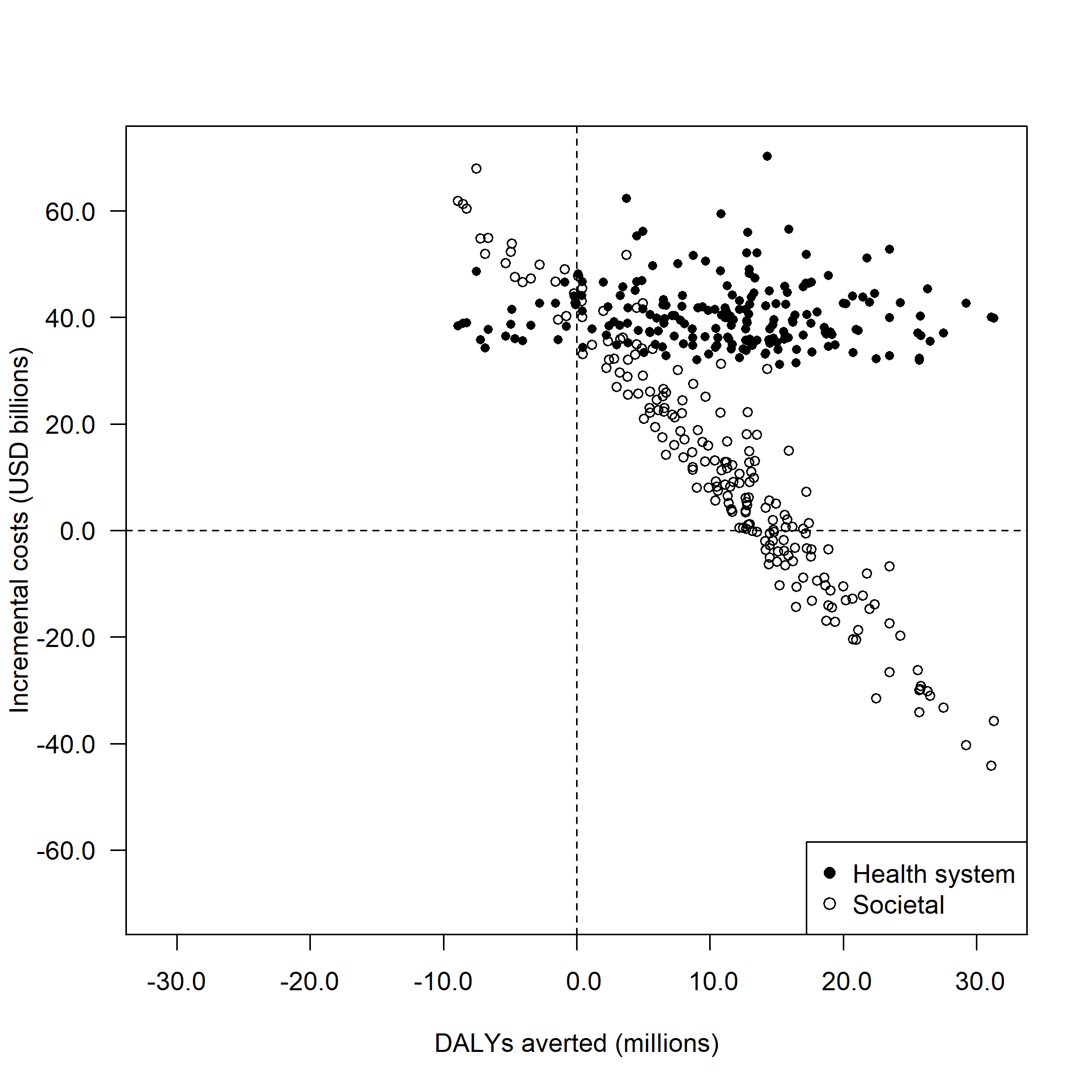

**Figure S 6**	**Cost-effectiveness plane of incremental costs and disability-adjusted life years (DALYs) averted across 200 probabilistic simulations in the *Main analysis*, by analytic perspective.**

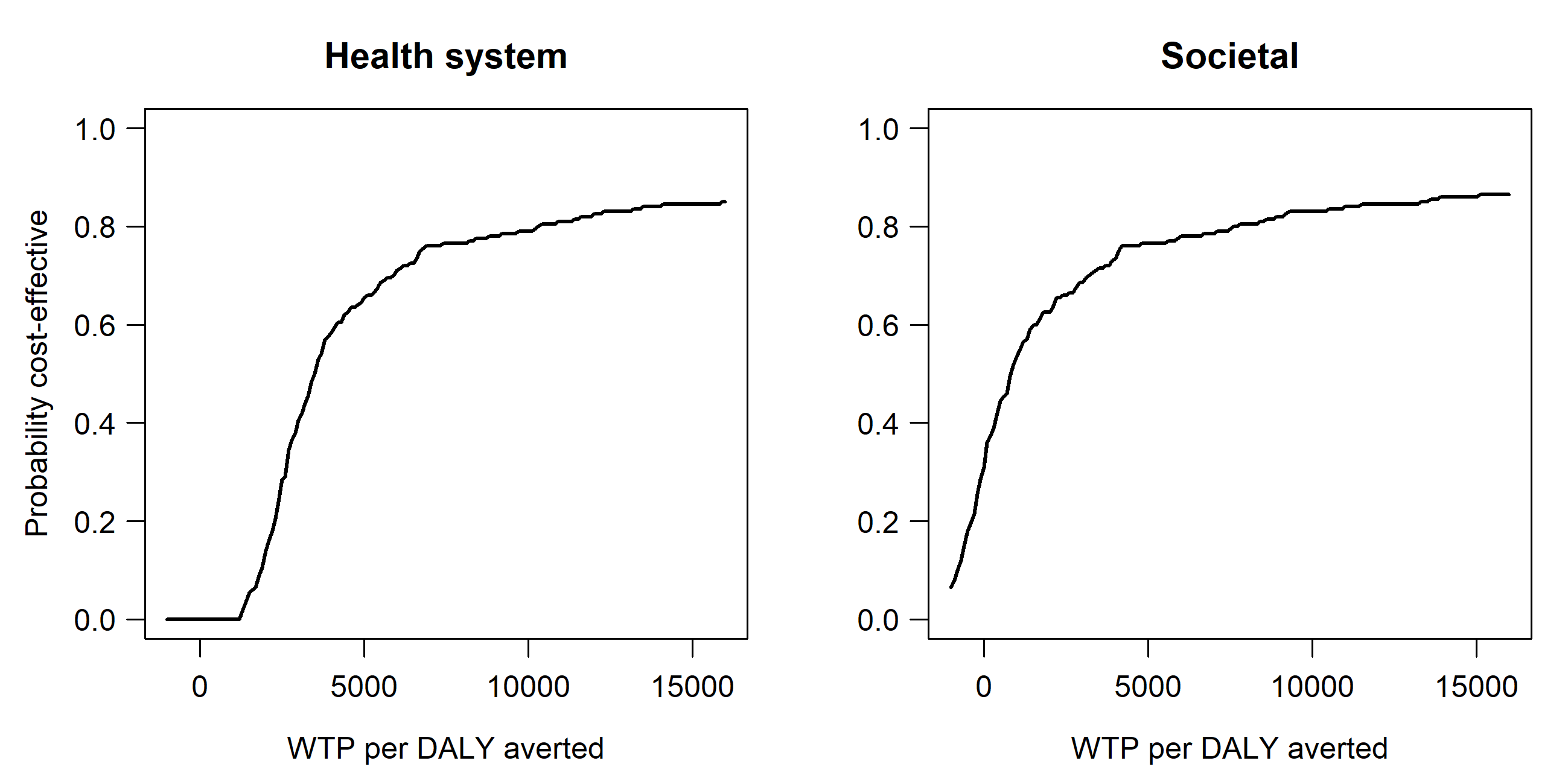

**Figure S 7**	**Cost-effectiveness acceptability curves by willingness-to-pay threshold in the *Main analysis*, stratified by analytic perspective.**

Note: Costs from the health system perspective include vaccination costs, TB testing and treatment costs, and antiretroviral treatment costs. Costs from the societal perspective include health system perspective costs, as well as patient non-medical costs and productivity losses. DALY = disability-adjusted life year; WTP = willingness-to-pay.

**Table S 8	Discounted costs, DALYs averted, and cost-effectiveness of VPM1002 candidate TB vaccine by analytic scenario, assuming $0.50 vaccine price per dose.**

| **Scenario** | **Health system perspective^a^ incremental cost (USD billions)** | **Societal perspective^b^ incremental cost (USD billions)** | **DALYs averted (millions)** | **Health system cost (USD) per DALY averted** | **Societal cost (USD) per DALY averted** |
| --- | --- | --- | --- | --- | --- |
| **Main analysis** | 38.2 (30 to 53.7) | 9.1 (-34 to 52.5) | 11.0 (-6.9 to 26.4) | 3466 (1259 to dominated) | 827 (dominant to dominated) |
| **Alternative scenarios: Varying duration of protection and introduction year scenarios** | | | | | |
| **10 years duration of protection** | 32.6 (25.2 to 45.9) | -17.3 (-87.7 to 61.9) | 18.9 (-12.3 to 44.5) | 1724 (616 to dominated) | dominant (dominant to dominated) |
| **All introduce in 2027** | 40.8 (31.8 to 56.6) | 0.8 (-56.9 to 61.9) | 15.1 (-9.3 to 35.8) | 2710 (999 to dominated) | 50.0 (dominant to dominated) |
| **Early adopter** | 39.8 (30.9 to 55.3) | 0.4 (-57.1 to 60.5) | 14.4 (-8.7 to 34.4) | 2770 (1036 to dominated) | 27.5 (dominant to dominated) |
| **Alternative scenarios: Varying efficacy scenarios** | | | | | |
| **Per protocol** | 38 (30.4 to 52.6) | 0.6 (-44.8 to 48.3) | 14.1 (-1.9 to 30.3) | 2686 (1186 to dominated) | 40 (dominant to dominated) |
| **Prevents PTB-only** | 38.4 (30.1 to 53.5) | 16.8 (-28.0 to 71.2) | 8.2 (-11.6 to 23.2) | 4703 (1463 to dominated) | 2057 (dominant to dominated) |
| **Alternative scenarios: Varying age group and varying who the vaccine works in** | | | | | |
| **Young** | 9.3 (7.8 to 11.5) | -12.0 (-40.3 to 25.1) | 8.3 (-6.6 to 19.2) | 1130 (446 to dominated) | dominant (dominant to dominated) |
| **Uninfected** | 40.1 (32 to 56.6) | 12.5 (-19.5 to 54.5) | 10.6 (-1.4 to 20.5) | 3781 (1690 to dominated) | 1178 (dominant to dominated) |

^a^ Costs from the health system perspective include vaccination costs, TB testing and treatment costs, and antiretroviral treatment costs.

^b^ Costs from the societal perspective include health system perspective costs, as well as patient non-medical costs and productivity losses.

Note: Introduction of VPM1002 is considered ‘dominant’ if it is both less costly and more effective (in this analysis, greater DALYs averted) compared to no-new-vaccine introduction, whereas introduction of VPM1002 is considered ‘dominated’ if it is both more costly and less effective compared to no-new-vaccine introduction. DALY = disability-adjusted life year.

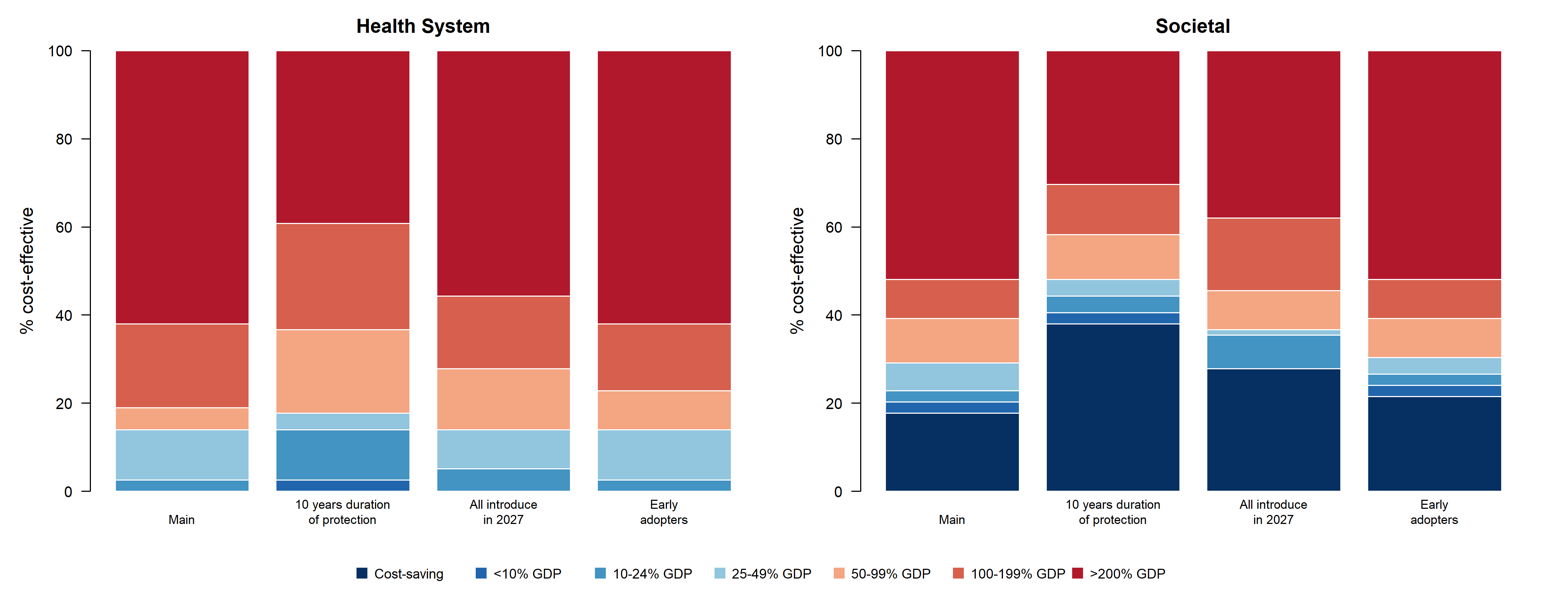

**Figure S 8	Percentage of countries where vaccination may be cost-effective compared to percentage of gross domestic product per capita thresholds, comparing health system and societal perspectives, for the *Main analysis* compared to 10 years duration of protection, all countries introducing in 2027, and early adopter countries, assuming $0.50 vaccine price per dose.**

Note: Countries include 79 LMICs analysed. GDP per capita estimates from 2023. GDP, gross domestic product per capita; LMIC, low- and middle-income country.

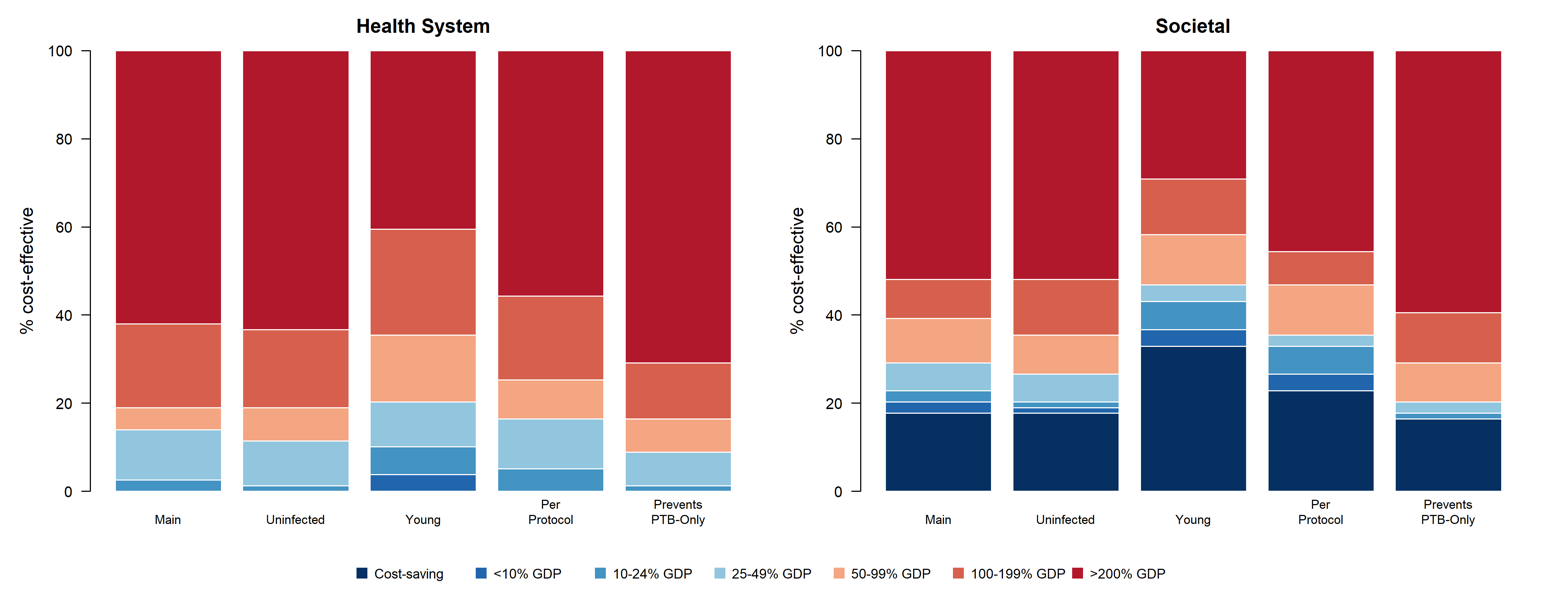

**Figure S 9	Percentage of countries where vaccination may be cost-effective compared to percentage of gross domestic product per capita thresholds, comparing health system and societal perspectives, by vaccine efficacy scenario, assuming $0.50 vaccine price per dose.**

Note: Countries include 79 LMICs analysed. GDP per capita estimates from 2023. GDP, gross domestic product per capita; LMIC, low- and middle-income country.

**Table S 9	Discounted costs, DALYs averted, and cost-effectiveness of VPM1002 candidate TB vaccine by analytic scenario, assuming $1.00 vaccine price per dose**

| **Scenario** | **Health system perspective^a^ incremental cost**  **(USD billions)** | **Societal perspective^b^ incremental cost (USD billions)** | **DALYs averted (millions)** | **Health system cost (USD) per DALY averted** | **Societal cost (USD) per DALY averted** |
| --- | --- | --- | --- | --- | --- |
| **Main analysis** | 42.8 (34.6 to 58.3) | 13.7 (-29.3 to 57.1) | 11.0 (-6.9 to 26.4) | 3886 (1427 to dominated) | 1246 (dominant to dominated) |
| **Alternative scenarios: Varying duration of protection and introduction year scenarios** | | | | | |
| **10 years duration of protection** | 36.6 (29.2 to 49.9) | -13.2 (-83.7 to 65.9) | 18.9 (-12.3 to 44.5) | 1936 (703 to dominated) | dominant (dominant to dominated) |
| **All introduce in 2027** | 45.8 (36.8 to 61.5) | 5.7 (-52.0 to 66.9) | 15.1 (-9.3 to 35.8) | 3040 (1133 to dominated) | 380 (dominant to dominated) |
| **Early adopter** | 44.6 (35.7 to 60.2) | 5.2 (-52.2 to 65.4) | 14.4 (-8.7 to 34.4) | 3107 (1177 to dominated) | 365 (dominant to dominated) |
| **Alternative scenarios: Varying efficacy scenarios** | | | | | |
| **Per protocol** | 42.6 (35.0 to 57.2) | 5.2 (-40.2 to 52.9) | 14.1 (-1.9 to 30.3) | 3012 (1330 to dominated) | 367 (dominant to dominated) |
| **Prevents PTB-only** | 43.0 (34.7 to 58.1) | 21.4 (-23.3 to 75.8) | 8.2 (-11.6 to 23.2) | 5269 (1642 to dominated) | 2623 (dominant to dominated) |
| **Alternative scenarios: Varying age group and varying who the vaccine works in** | | | | | |
| **Young** | 10.5 (9.0 to 12.7) | -10.8 (-39.2 to 26.3) | 8.3 (-6.6 to 19.2) | 1271 (506 to dominated) | dominant (dominant to dominated) |
| **Uninfected** | 44.9 (36.8 to 61.4) | 17.3 (-14.7 to 59.3) | 10.6 (-1.4 to 20.5) | 4238 (1906 to dominated) | 1635 (dominant to dominated) |

^a^ Costs from the health system perspective include vaccination costs, TB testing and treatment costs, and antiretroviral treatment costs.

^b^ Costs from the societal perspective include health system perspective costs, as well as patient non-medical costs and productivity losses.

Note: Introduction of VPM1002 is considered ‘dominant’ if it is both less costly and more effective (in this analysis, greater DALYs averted) compared to no-new-vaccine introduction, whereas introduction of VPM1002 is considered ‘dominated’ if it is both more costly and less effective compared to no-new-vaccine introduction. DALY = disability-adjusted life year.

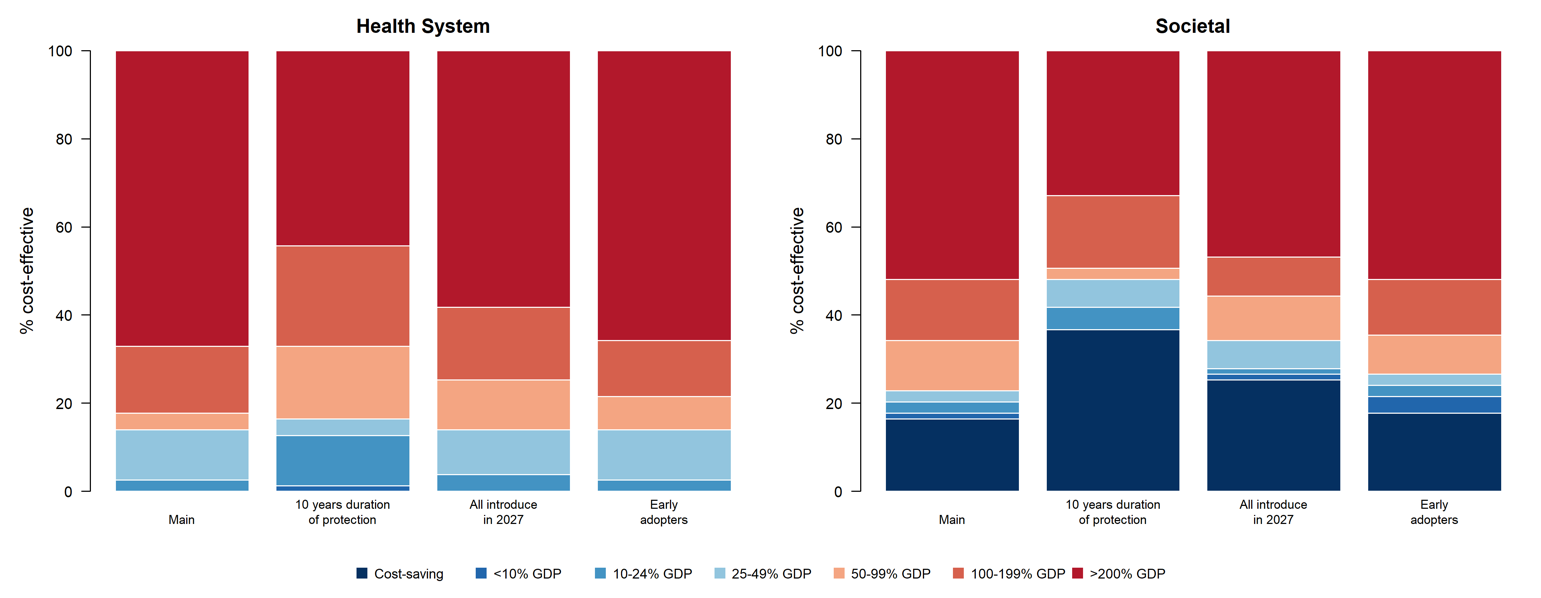

**Figure S 10	Percentage of countries where vaccination may be cost-effective compared to percentage of gross domestic product per capita thresholds, comparing health system and societal perspectives, for the main analysis compared to 10 years duration of protection, all countries introducing in 2027, and early adopter countries, assuming $1.00 vaccine price per dose.**

Note: Countries include 79 LMICs analysed. GDP per capita estimates from 2023. GDP, gross domestic product per capita; LMIC, low- and middle-income country.

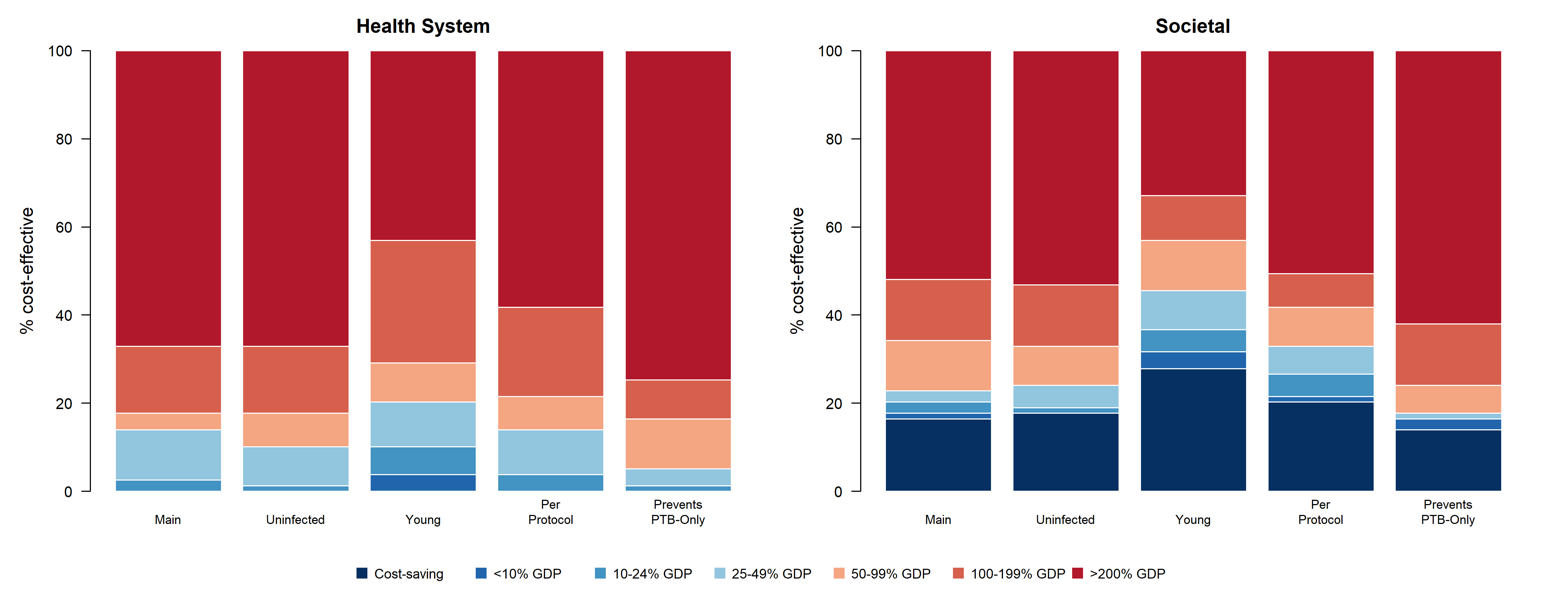

**Figure S 11	Percentage of countries where vaccination may be cost-effective compared to percentage of gross domestic product per capita thresholds, comparing health system and societal perspectives, by vaccine efficacy scenario, assuming $1.00 vaccine price per dose.**

Note: Countries include 79 LMICs analysed. GDP per capita estimates from 2023. GDP, gross domestic product per capita; LMIC, low- and middle-income country.
